# Efficacy and Safety of Pharmacologic Therapies in HFpEF: A Systematic Review and Network Meta-Analysis

**DOI:** 10.1101/2025.09.17.25335973

**Authors:** Maaz Nazir Armani, Abhishek Mehan, Dhruvilkumar patel, Yashas Maragowdanahalli Somegowda, Kalyani Gadidesi, Mahveen Nishat, Nabiha Muneer, Rashika Reddy Ananthula, Mariya Hussain Bingursain, Swetcha Reddy Komati Reddy, Rakhshanda khan, Harshawardhan Dhanraj Ramteke, Manish juneja

## Abstract

**Introduction:** Heart failure with preserved ejection fraction (HFpEF) represents nearly half of all heart failure cases worldwide and remains a major cause of morbidity and mortality, especially in older adults. Despite advances in pharmacotherapy, no single intervention has consistently improved survival outcomes in this heterogeneous population. Multiple drug classes, including SGLT2 inhibitors, angiotensin receptor-neprilysin inhibitors (ARNI), mineralocorticoid receptor antagonists (MRAs), and other agents, have demonstrated varying efficacy across clinical endpoints. A network meta-analysis allows comprehensive comparison of direct and indirect evidence across therapies to clarify their relative effectiveness and safety.

**Methods:** We conducted a systematic review and network meta-analysis of randomized controlled trials (RCTs) in patients with HFpEF. Databases including PubMed, Embase, Cochrane CENTRAL, and Web of Science were searched up to September 2025. Eligible studies compared pharmacologic therapies with placebo or active comparators and reported outcomes including all-cause mortality, cardiovascular mortality, rehospitalization, functional capacity, quality of life (KCCQ), left ventricular ejection fraction (LVEF), NT-proBNP, and safety endpoints such as hyperkalemia, hypotension, and renal outcomes. Data extraction was performed independently, and risk of bias was assessed using RoB 2.0. Random-effects network meta-analysis was applied, and results were expressed as odds ratios (OR), mean differences (MD), or risk ratios (RR) with 95% credible intervals (CrI).

**Results:** From 968 studies screened, 37 RCTs with 65,068 patients (34,178 men and 30,447 women; mean age 70.6 ± 8 years, mean follow-up 16.1 ± 3 months) were included. Of these, 33,986 received active treatment and 30,289 were assigned to placebo or standard therapy. Across the network, all-cause mortality was not significantly reduced by any drug, with odds ratios overlapping unity; beta-blockers (OR 0.88, 95% CrI 0.17–4.56) and sacubitril/valsartan (OR 0.99, 95% CrI 0.18–5.54) showed neutrality, while vericiguat trended toward harm (OR 3.95, 95% CrI 0.64–24.8). For cardiovascular mortality, sacubitril/valsartan (OR 0.62, 95% CI –1.42 to 0.17) and finerenone (OR 0.93, 95% CI 0.81–1.05) demonstrated benefit, while vericiguat increased risk (OR 1.67, 95% CI 0.58–2.75). Rehospitalization was significantly reduced by sacubitril/valsartan (RR –0.28, 95% CI –0.49 to – 0.08) and finerenone (RR –0.21, 95% CI –0.30 to –0.13), with dapagliflozin and tirzepatide showing additional favorable effects. Functional outcomes improved modestly, with treatment groups achieving a mean 6-minute walk distance of 248.6 ± 129 cm compared with 240.5 ± 130.8 cm in controls. Quality of life (KCCQ scores) improved with dapagliflozin (MD +1.60), tirzepatide (MD +9.90), pirfenidone (MD +11.5), and semaglutide (MD +7.0). Biomarker analysis showed NT-proBNP reductions with sacubitril/valsartan (MD –188.5) and spironolactone (MD –200), whereas vericiguat increased levels. LVEF changes were neutral across most agents. Safety analysis revealed increased hyperkalemia with finerenone, while sacubitril/valsartan and MRAs were associated with higher risks of hypotension and renal impairment, underscoring the need for careful clinical monitoring.

**Conclusion:** This network meta-analysis demonstrates that while no pharmacologic therapy significantly reduces all-cause mortality in HFpEF, certain agents, notably sacubitril/valsartan, finerenone, and SGLT2 inhibitors, improve morbidity outcomes, reduce rehospitalizations, and enhance quality of life. Safety concerns, particularly hyperkalemia and renal dysfunction with MRAs, highlight the importance of careful patient selection and monitoring. These findings support the integration of multimodal therapy for morbidity reduction in HFpEF, while emphasizing the urgent need for future trials to address survival outcomes.

## Introduction

Heart failure (HF) is a major global health burden, affecting an estimated >64 million people and driving substantial morbidity, hospitalizations, and costs worldwide. Heart failure with preserved ejection fraction (HFpEF) now accounts for roughly half of all HF, and its prevalence is rising with population aging and multimorbidity. Patients are often older, more frequently women, and commonly have obesity, hypertension, diabetes, atrial fibrillation, and chronic kidney disease—factors that contribute to frequent decompensations and poor quality of life despite “preserved” systolic function. Unlike HFrEF, evidence-based disease-modifying therapies for HFpEF have historically been elusive, leaving a large and growing care gap. [1–3].

HFpEF pathophysiology is heterogeneous and systemic, encompassing coronary microvascular/endothelial dysfunction, low-grade inflammation, myocardial fibrosis, ventricular–vascular stiffening, and impaired diastolic reserve. The influential paradigm proposed by Paulus and Tschöpe posits that comorbidity-driven systemic inflammation induces coronary microvascular endothelial dysfunction, nitric oxide–cGMP–PKG signaling downregulation, and increased cardiomyocyte stiffness—mechanistic hallmarks now widely cited to explain HFpEF’s limited response to traditional neurohormonal therapies [4]. Contemporary reviews further highlight inflammatory signaling, extracardiac organ interplay, and phenotypic diversity as key barriers to “one-size-fits-all” therapy [5].

Clinically, diuretics remain foundational for decongestion yet do not improve survival. Multiple attempts to repurpose renin–angiotensin–aldosterone system (RAAS) blockade have yielded neutral or modest findings: ACE inhibitors (PEP-CHF, perindopril) and ARBs (CHARM-Preserved, candesartan; I-PRESERVE, irbesartan) showed at best small reductions in HF hospitalization, without consistent mortality benefit [5–7]. These results helped establish that classic RAAS blockade alone is insufficient to alter HFpEF’s trajectory.

Mineralocorticoid receptor antagonism has been evaluated most prominently in TOPCAT (spironolactone), which did not significantly reduce its primary composite outcome, though regional signal heterogeneity and a modest reduction in HF hospitalization in some analyses were observed [8]. Non-steroidal MRAs (e.g., finerenone) are under active investigation but lack definitive outcomes in HFpEF. In parallel, angiotensin receptor–neprilysin inhibition (ARNI) with sacubitril/valsartan in PARAGON-HF narrowly missed statistical significance for its composite of total HF hospitalizations and cardiovascular (CV) death; post-hoc subgroup findings (e.g., women, lower EF within the preserved range) suggest possible heterogeneity of treatment effect warranting further study [9].

The most consequential progress has come from sodium–glucose cotransporter-2 inhibitors (SGLT2i). EMPEROR-Preserved (empagliflozin) and DELIVER (dapagliflozin) demonstrated consistent reductions in the composite of CV death or HF worsening—benefits largely driven by fewer HF hospitalizations and observed irrespective of diabetes status. These trials also reported improvements in patient-reported outcomes, positioning SGLT2 inhibitors as the first broadly efficacious pharmacologic option across the HFpEF spectrum [10–12]. Shorter-term trials further showed clinically meaningful KCCQ gains with dapagliflozin, underscoring symptomatic/functional benefits alongside event reduction [12]. Persisting questions include the relative magnitude of benefit versus other classes, mortality effects, and performance across phenotypes (e.g., obesity- inflammatory or AF-predominant HFpEF).

By contrast, several alternative pathways have delivered neutral results. Organic nitrates (NEAT-HFpEF) were associated with reduced daily activity and no QoL gains; phosphodiesterase-5 inhibition (RELAX) did not improve exercise capacity or clinical status; soluble guanylate cyclase stimulation (VITALITY-HFpEF, vericiguat) failed to improve KCCQ physical limitation scores after recent decompensation [13–15]. Novel antifibrotic therapy with pirfenidone (PIROUETTE) reduced cardiac extracellular volume (ECV) on CMR over 52 weeks—an encouraging mechanistic signal—yet clinical outcome benefits remain unproven and require larger trials [16].

This heterogeneous, partly conflicting evidence base leaves clinicians with multiple options—SGLT2i, ARNI, MRAs, ACEi/ARBs, β-blockers (for rate control/AF/hypertension), diuretics for symptoms, and emerging antifibrotic or NO–cGMP pathway agents—without clear head-to-head comparisons to guide which therapy should be prioritized for which patient. Traditional pairwise meta-analyses synthesize direct comparisons versus placebo/standard care but cannot simultaneously compare multiple active treatments or derive relative hierarchies when head-to-head trials are absent. In contrast, a network meta-analysis (NMA) can integrate direct and indirect evidence from randomized trials across drug classes to (1) estimate comparative efficacy and safety, (2) generate treatment rankings for clinically important outcomes (HF hospitalization, CV death, composite endpoints), (3) incorporate patient-centered endpoints (KCCQ, 6MWT), and (4) explore effect modifiers (sex, EF spectrum, comorbidities). Given HFpEF’s diverse biology and variable treatment responses, an NMA is uniquely suited to clarify therapeutic positioning, inform guidelines, and highlight evidence gaps for future trials [3,9–12].

## Methods

### Literature Search

A comprehensive literature search will be conducted in PubMed/MEDLINE, Embase, Cochrane CENTRAL, and Web of Science from database inception to the present. The strategy will combine controlled vocabulary (e.g., MeSH, Emtree) and free-text terms related to “heart failure with preserved ejection fraction” and drug therapies including SGLT2 inhibitors, angiotensin receptor–neprilysin inhibitors, mineralocorticoid receptor antagonists, ACE inhibitors, ARBs, β-blockers, diuretics, and emerging agents (pirfenidone, vericiguat, nitrates, PDE-5 inhibitors, digoxin, ivabradine). Only randomized controlled trials in adults will be included. References of eligible articles and relevant reviews will be screened to ensure completeness. It followed PRISMA guidelines [17]. It is registered in Prospero with number CRD420251149394.

### Study Selection and Data Extraction

Two reviewers will independently screen all retrieved records using titles and abstracts, followed by full-text assessment for eligibility. Discrepancies will be resolved through discussion or adjudication by a third reviewer. Eligible studies will include randomized controlled trials enrolling adults with HFpEF, defined as left ventricular ejection fraction ≥50% or as specified by trial authors. Interventions of interest will comprise SGLT2 inhibitors, ARNIs, MRAs, ACE inhibitors, ARBs, β-blockers, diuretics, and other emerging agents, compared with placebo or active controls.

Data will be extracted using a standardized form, capturing trial characteristics (author, year, country, sample size, follow-up), participant demographics (age, sex, comorbidities, baseline EF), interventions and comparators (drug, dose, duration), and outcomes. Primary outcomes will include all-cause mortality, cardiovascular mortality, and HF hospitalization. Secondary outcomes will encompass composite endpoints, quality of life measures (KCCQ, MLHFQ), functional capacity (6MWT, peak VO), biomarkers (NT-proBNP, renal indices), echocardiographic parameters (E/e′, GLS, LA volume, LV mass), and safety events (hyperkalemia, renal dysfunction, hypotension, infection, hepatotoxicity).

Where multiple reports of the same trial exist, the most complete dataset will be used. Extracted data will be cross-checked, and trial investigators will be contacted if clarification is required.

### Risk of Bias

Risk of bias will be independently assessed by two reviewers using the Cochrane Risk of Bias 2.0 tool for randomized controlled trials [18]. Domains evaluated will include randomization, deviations from intended interventions, missing data, outcome measurement, and reporting bias. Disagreements will be resolved through consensus or third-party adjudication.

### Statistical Analysis

We will perform a random effects network meta analysis (NMA) to compare the relative efficacy and safety of all included interventions. Pairwise meta analyses will first be conducted for direct comparisons, followed by a Bayesian framework NMA to integrate direct and indirect evidence. Effect estimates will be expressed as risk ratios (RRs) with 95% credible intervals (CrI) for dichotomous outcomes (all-cause mortality, cardiovascular mortality, HF hospitalization) and mean differences (MDs) or standardized mean differences (SMDs) for continuous outcomes (KCCQ scores, 6MWT, NT proBNP, echocardiographic indices). Surface under the cumulative ranking curve (SUCRA) values will be calculated to rank interventions. Statistical heterogeneity will be assessed using the I² statistic, while global and local inconsistency will be evaluated through node splitting and design by treatment interaction models. Publication bias will be examined via comparison adjusted funnel plots. All analyses will be conducted using R (netmeta, gemtc) and Stata software.

## Results

### Demographics

Out of 968 studies initially identified, 37 randomized controlled trials (RCTs) [19–54] with a total of 65,068 participants were included in the final analysis. Of these, 34,178 were male and 30,447 were female, with a mean age of 70.6 years (SD 8.0). The average follow-up duration was 16.1 months (SD 3.0). Across trials, the mean left ventricular ejection fraction (LVEF) was 56.1% (SD 8.5) in the treatment arms and 56.2% (SD 9.3) in the control arms. Functional capacity, as assessed by the six-minute walk test (6MWT), averaged 248.6 m (SD 129.0) in treatment groups and 240.5 m (SD 130.8) in control groups. In total, 33,986 patients were randomized to active treatment and 30,289 patients to control groups. The interventions evaluated included β-blockers (n=5,177; bisoprolol 276, carvedilol 746, nebivolol 57), SGLT2 inhibitors (dapagliflozin 3,145; empagliflozin 3,189; canagliflozin 222), ARNI (sacubitril/valsartan 4,307), MRAs (spironolactone 3,518; finerenone 11,801), ARBs (valsartan 4,395; olmesartan 574), as well as novel and emerging therapies including dapagliflozin plus spironolactone (105), pirfenidone (47), vericiguat (264), nitrates such as isosorbide mononitrate (110) and potassium nitrate (77), ivabradine (95), and potassium chloride (74). Placebo groups comprised 25,159 patients. Collectively, these trials represent the broad spectrum of pharmacological strategies explored for HFpEF, ranging from conventional neurohormonal modulators to emerging antifibrotic and metabolic therapies. Table S1.

### Network Plot and Forest Plot for the All-Cause Mortality

Figure 2 illustrates the network of interventions and the corresponding forest plot for all-cause mortality compared with placebo. Across the included trials, none of the pharmacological agents demonstrated a statistically significant reduction in mortality, as all 95% credible intervals crossed unity. Beta-blockers overall showed an odds ratio (OR) of 0.88 (95% CrI 0.17–4.56), with bisoprolol (OR 2.26, 95% CrI 0.32–16.1) and carvedilol (OR 0.88, 95% CrI 0.15–5.04) demonstrating wide intervals and substantial uncertainty. Empagliflozin (OR 0.99, 95% CrI 0.02–48.3) and finerenone (OR 1.29, 95% CrI 0.50–3.34) were neutral, while spironolactone (OR 1.67, 95% CrI 0.31–8.87) similarly showed no significant effect. Ivabradine (OR 3.58, 95% CrI 0.22–144.0), olmesartan (OR 1.16, 95% CrI 0.22–6.19), and vericiguat (OR 3.95, 95% CrI 0.64–24.8) also failed to demonstrate consistent benefit. Newer agents such as semaglutide (OR 0.99, 95% CrI 0.18–5.54) and tirzepatide (OR 0.51, 95% CrI 0.09–2.98) showed numerically lower odds of mortality, but the wide confidence intervals preclude definitive conclusions. Overall, these findings underscore that despite a broad spectrum of pharmacological strategies, no therapy to date has conferred a clear mortality benefit in HFpEF. Figure S1 and Table S2.

**Figure 1.**
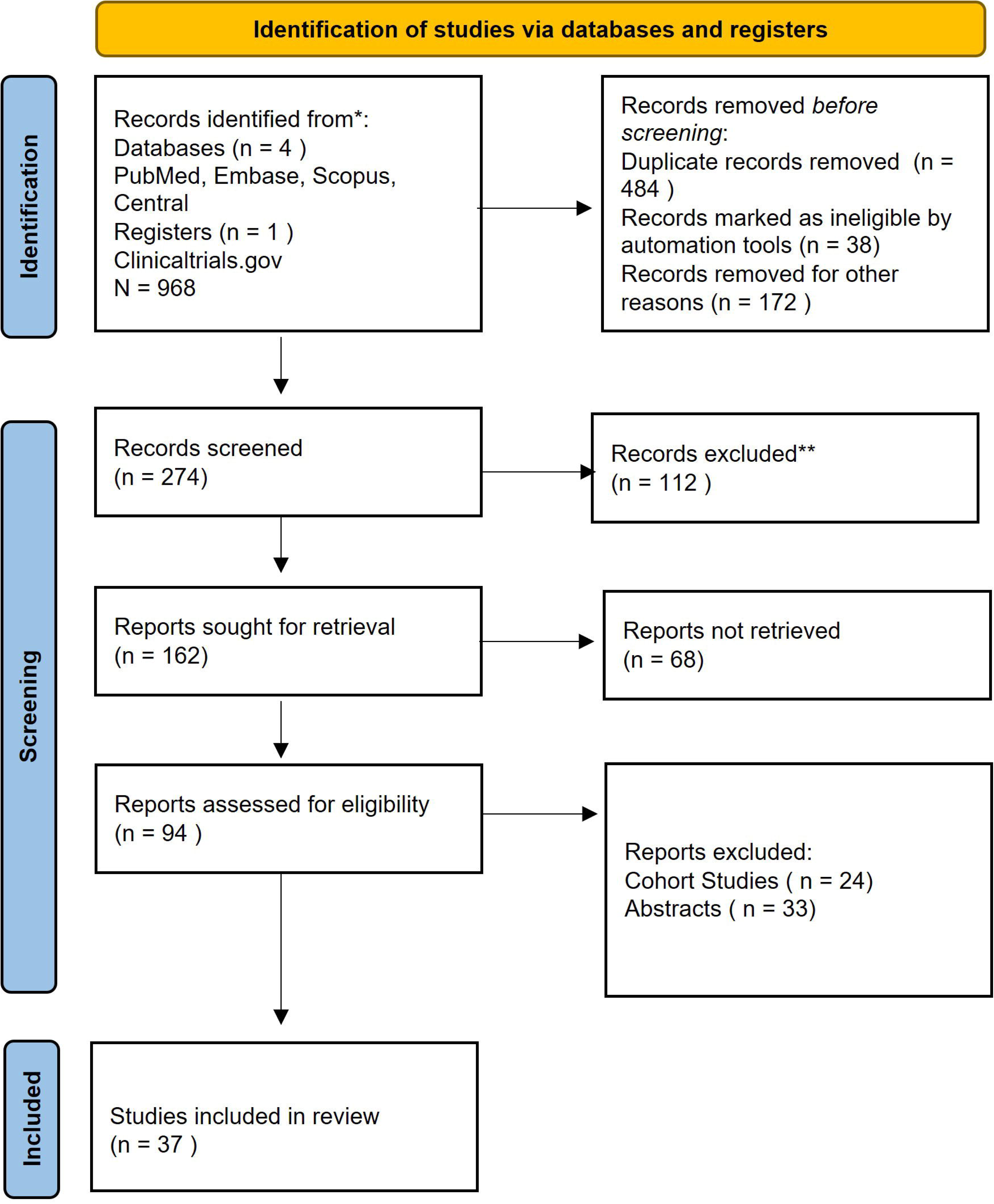
PRISMA Flow Diagram.

**Figure 2.**
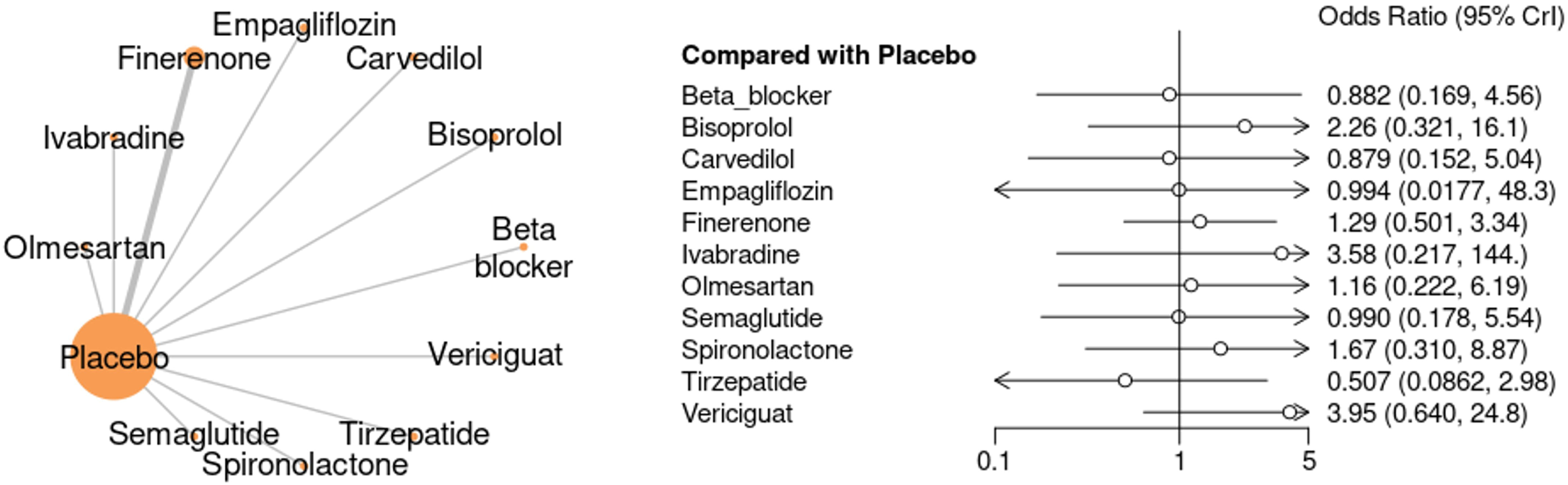
Network Plot and Forest plot for the All-Cause Mortality in the study.

Figure 3 summarizes the effects of individual pharmacological interventions on all-cause mortality compared with control. Beta-blockers as a class showed a non-significant reduction in mortality (log OR –0.12, 95% CI – 0.30 to 0.05), with bisoprolol (log OR 0.79, 95% CI –0.33 to 1.91) and carvedilol (log OR –0.13, 95% CI –0.82 to 0.55) both neutral. Empagliflozin (log OR 1.01, 95% CI –2.77 to 2.79) showed no mortality benefit. Finerenone displayed a pooled effect of log OR 0.26 (95% CI –0.41 to 0.92), again non-significant with high heterogeneity across trials. Ivabradine (log OR 1.00, 95% CI –1.29 to 3.28) and olmesartan (log OR 0.15, 95% CI –0.17 to 0.47) were neutral. Sacubitril/valsartan demonstrated heterogeneous results, ranging from benefit in some trials (e.g., Rambarat et al. 2025, log OR –0.62) to potential harm in others (Vadyanathan et al. 2020, log OR 1.28), yielding an overall non-significant pooled effect (log OR –0.01, 95% CI –0.92 to 0.91). Semaglutide (log OR 0.00, 95% CI –0.54 to 0.54) showed no effect, while spironolactone was associated with a modest but significant increased risk (log OR 0.51, 95% CI 0.11–0.90). Tirzepatide (log OR –0.67, 95% CI –1.41 to 0.08) showed a non-significant trend toward benefit, whereas vericiguat (log OR 1.34, 95% CI 0.48–2.19) suggested possible harm. The overall pooled estimate across all interventions was neutral (log OR 0.18, 95% CI –0.13 to 0.49), indicating that no therapy demonstrated a consistent mortality benefit in HFpEF.

**Figure 3.**
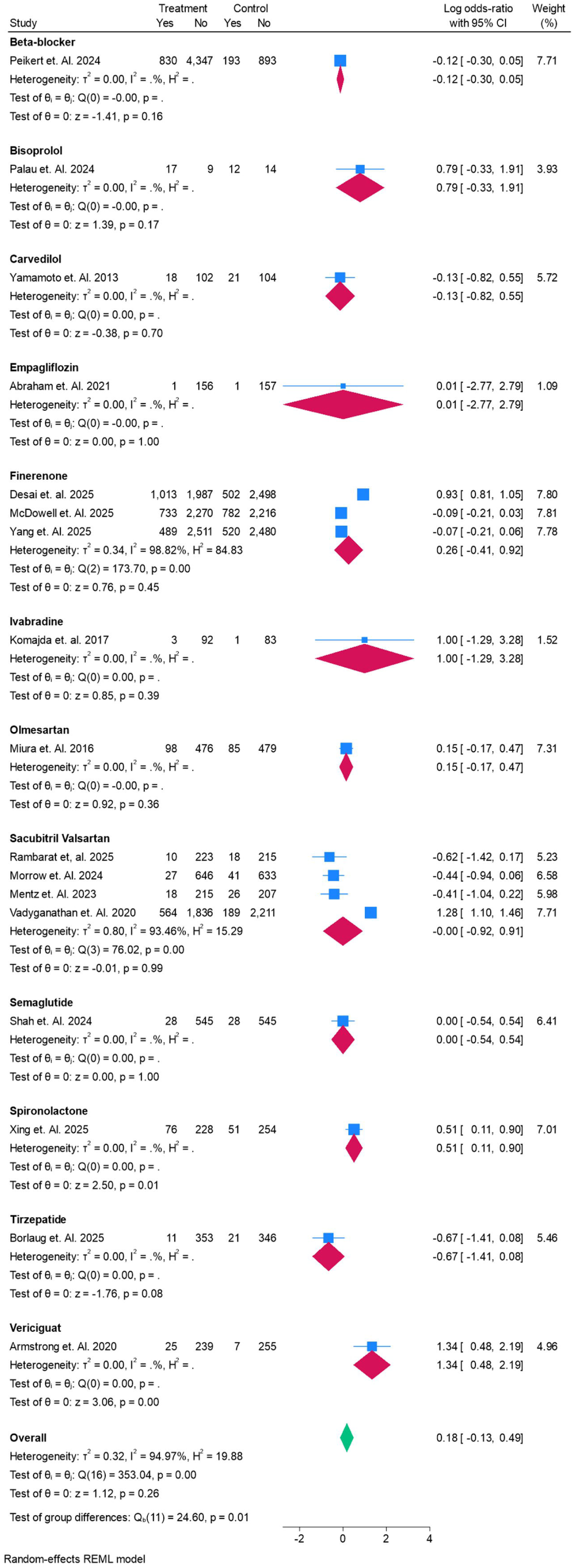
Forest plot for the All-Cause Mortality in the study.

### Cardiovascular Mortality

Figure 4 presents the network of interventions and the comparative effects of pharmacological therapies versus placebo on cardiovascular mortality. Overall, none of the tested agents demonstrated a statistically significant reduction in cardiovascular death, as all 95% credible intervals crossed unity. Beta-blockers as a class showed a trend toward reduced risk (OR 0.74, 95% CrI 0.11–4.94), though this was not statistically significant. Among individual agents, bisoprolol (OR 1.63, 95% CrI 0.19–14.1), carvedilol (OR 1.20, 95% CrI 0.14–10.5), finerenone (OR 1.27, 95% CrI 0.43–3.79), ivabradine (OR 2.24, 95% CrI 0.10–109.0), spironolactone (OR 1.66, 95% CrI 0.24–11.4), and vericiguat (OR 5.74, 95% CrI 0.67–53.8) were neutral or trended toward harm. Empagliflozin (OR 2.51, 95% CrI 0.10–116.0) also showed no benefit, with wide uncertainty. Tirzepatide (OR 0.42, 95% CrI 0.05–3.24) was the only agent suggesting a possible favorable trend, but the wide interval precludes firm conclusions. Collectively, these findings indicate that no pharmacological intervention has yet demonstrated a consistent cardiovascular mortality benefit in HFpEF. Figure S2 and Table S3

**Figure 4.**
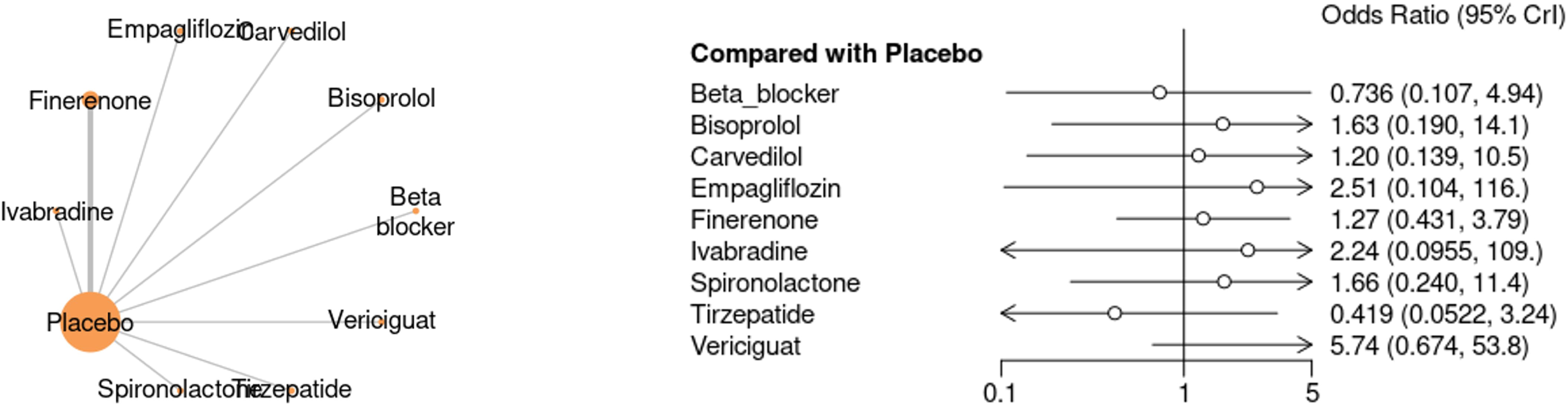
Network Plot and Forest plot for the Cardiovascular Mortality in the study.

Figure 5 displays the pooled effects of individual pharmacological interventions on cardiovascular mortality. Beta-blockers demonstrated a statistically significant reduction in risk (log OR –0.31, 95% CI –0.53 to –0.08), largely driven by Peikert et al. (2024). However, subgroup analyses for bisoprolol (log OR 0.46, 95% CI –0.63 to 1.56) and carvedilol (log OR 0.19, 95% CI –0.86 to 1.23) were neutral. Empagliflozin (log OR 0.71, 95% CI –1.70 to 3.12) and finerenone (pooled log OR 0.24, 95% CI –0.44 to 0.92) did not reduce mortality. Ivabradine showed no benefit (log OR 0.58, 95% CI –1.84 to 3.00). Sacubitril/valsartan demonstrated a favorable signal with pooled log OR –0.60 (95% CI –1.02 to –0.19), indicating a significant reduction in cardiovascular mortality across studies. Spironolactone was associated with increased risk (log OR 0.51, 95% CI 0.11–0.90). Tirzepatide trended toward benefit (log OR –0.83, 95% CI –1.68 to 0.02), approaching statistical significance. By contrast, vericiguat was linked to increased cardiovascular mortality (log OR 1.67, 95% CI 0.58–2.75). The overall pooled estimate (log OR 0.05, 95% CI –0.31 to 0.41) was neutral, suggesting that while some agents, notably sacubitril/valsartan and beta-blockers, show promise, consistent cardiovascular mortality benefit remains unproven across drug classes.

**Figure 5.**
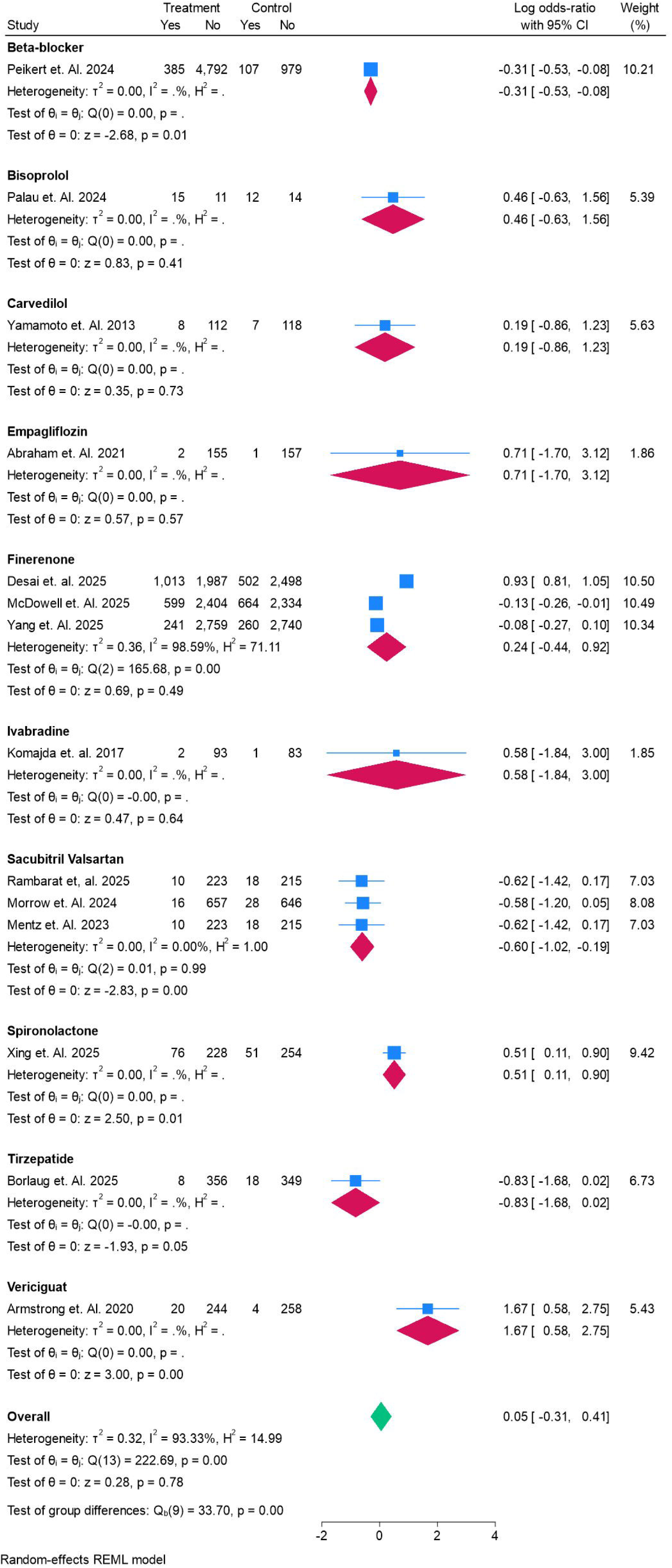
Forest plot for the Cardiovascular Mortality in the study.

### Rehospitalization

Figure 6 shows the network plot and forest plot comparing pharmacological therapies with placebo for the outcome of rehospitalization. Overall, none of the agents demonstrated a statistically significant reduction in rehospitalization, as all 95% credible intervals crossed unity. Bisoprolol was associated with a numerically higher risk (OR 4.83, 95% CrI 0.77–46.3), though this result was imprecise. Carvedilol (OR 0.76, 95% CrI 0.24–2.47) and empagliflozin (OR 0.68, 95% CrI 0.21–2.19) showed non-significant trends toward reducing rehospitalization, while finerenone (OR 0.81, 95% CrI 0.38–1.73) was neutral. Ivabradine (OR 1.21, 95% CrI 0.28–4.94) showed no benefit, and spironolactone (OR 0.64, 95% CrI 0.06–4.76) demonstrated a wide and inconclusive estimate. Tirzepatide (OR 0.53, 95% CrI 0.13–1.95) trended toward reduced rehospitalization, but with high uncertainty. Collectively, these findings highlight the absence of clear evidence that any pharmacological agent significantly prevents rehospitalization in HFpEF, though SGLT2 inhibitors and tirzepatide may warrant further investigation given favorable trends. Figure S3 and Table S4.

**Figure 6.**
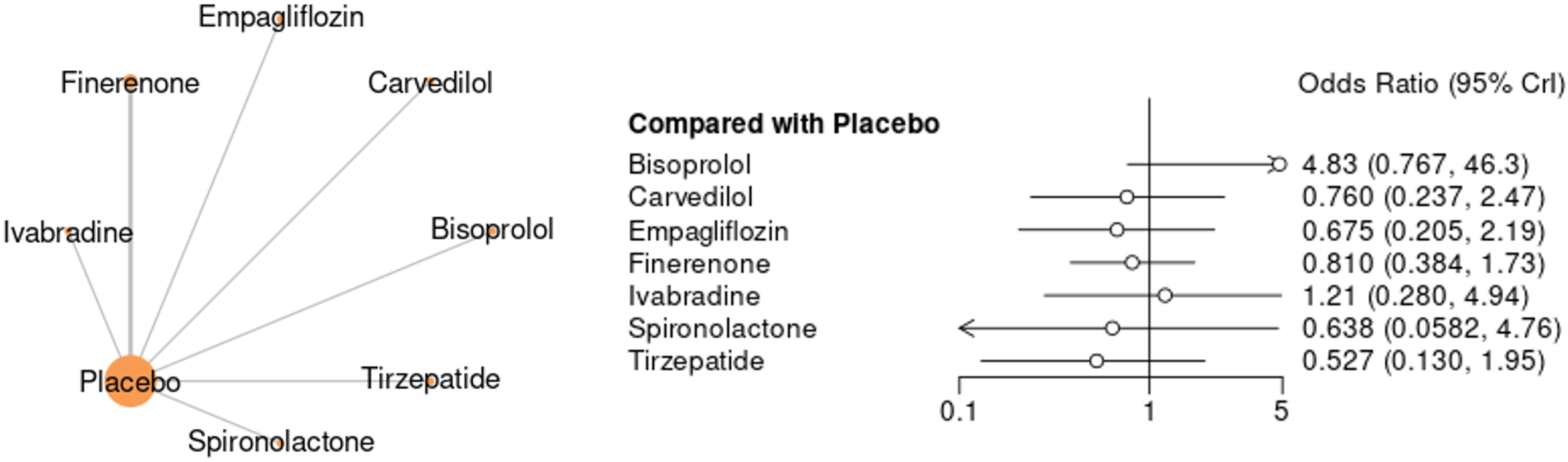
Network Plot and Forest plot for the Rehospitalization in the study.

Figure 7 presents the pooled effects of pharmacological therapies compared with control on rehospitalization in HFpEF. Overall, there was a statistically significant reduction in rehospitalization risk across treatments (pooled log OR –0.22, 95% CI –0.30 to –0.15). Among individual drugs, finerenone consistently demonstrated a significant benefit, with log OR –0.22 (95% CI –0.34 to –0.09) from McDowell et al. (2025) and –0.21 (95% CI –0.32 to –0.09) from Yang et al. (2025), yielding a robust pooled effect (–0.21, 95% CI –0.30 to –0.13). Sacubitril/valsartan also showed a favorable reduction (pooled log OR –0.28, 95% CI –0.49 to –0.08), with consistent findings across multiple trials. Empagliflozin demonstrated a non-significant trend toward fewer rehospitalizations (log OR –0.39, 95% CI –1.05 to 0.28), while tirzepatide also showed a non-significant trend in the same direction (–0.63, 95% CI –1.56 to 0.30). In contrast, bisoprolol (1.49, 95% CI –0.20 to 3.17) and ivabradine (0.14, 95% CI –0.89 to 1.18) did not reduce risk, while spironolactone (–0.41, 95% CI –2.20 to 1.39) had wide, inconclusive results. Collectively, these findings suggest that finerenone and sacubitril/valsartan provide consistent reductions in rehospitalization, while other agents demonstrate neutral or uncertain effects.

**Figure 7.**
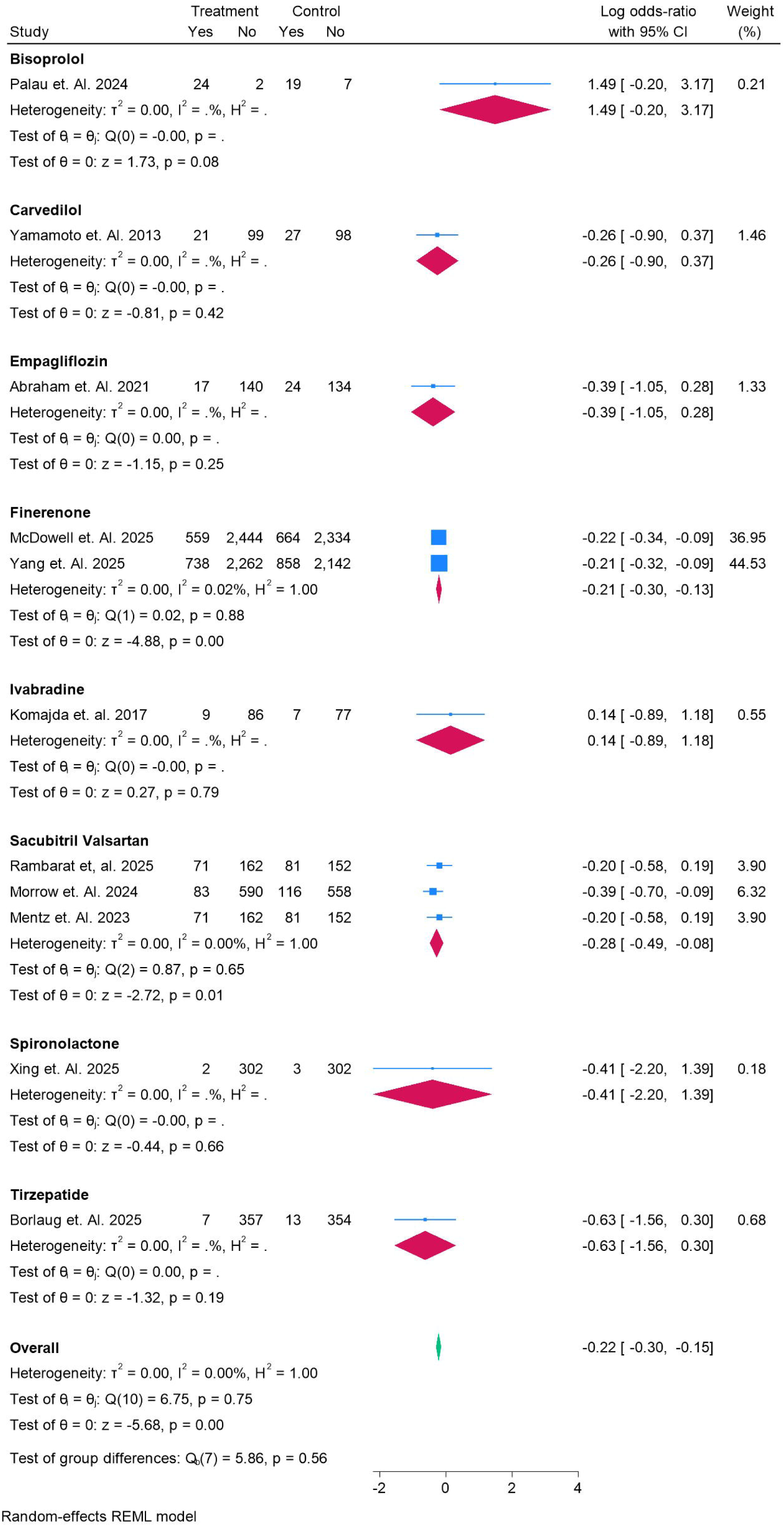
Forest plot for the Rehospitalization in the study.

### Quality of Life (KCCQ Scores)

Figure 8 illustrates the comparative effects of pharmacological interventions on patient-reported quality of life, measured by the Kansas City Cardiomyopathy Questionnaire (KCCQ) scores. Across the network, no intervention achieved a statistically significant improvement over placebo, as all 95% credible intervals crossed zero. Beta-blockers (MD 0.49, 95% CrI –13.1 to 14.0), empagliflozin (0.74, 95% CrI –7.14 to 8.59), and finerenone (–0.19, 95% CrI –13.8 to 13.6) were essentially neutral. SGLT2 inhibitors, including canagliflozin (3.34, 95% CrI –10.4 to 17.0), dapagliflozin (1.58, 95% CrI –11.8 to 15.2), and dapagliflozin plus spironolactone (–0.12, 95% CrI –19.2 to 19.2), showed no consistent benefit. Nebivolol (8.98, 95% CrI –4.82 to 22.6), pirfenidone (11.4, 95% CrI –3.09 to 25.8), semaglutide (7.02, 95% CrI –6.66 to 20.6), and tirzepatide (9.88, 95% CrI –3.85 to 23.6) all trended toward improved quality of life but with wide confidence intervals. By contrast, ISMN (–1.98, 95% CrI –15.7 to 11.7) and vericiguat (–3.65, 95% CrI –17.4 to 10.0) trended negatively. Overall, while some agents such as pirfenidone, nebivolol, semaglutide, and tirzepatide suggested potential for quality-of-life improvement, the lack of statistical significance highlights persistent uncertainty in patient-centered outcomes. Figure S4 and Table S5.

**Figure 8.**
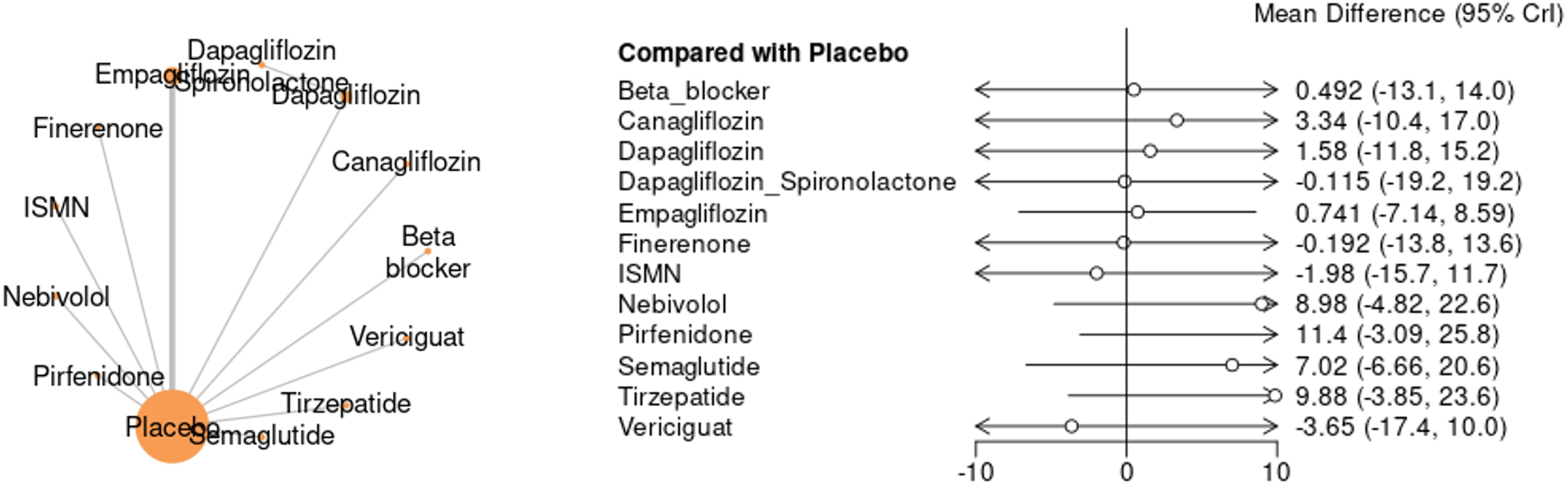
Network Plot and Forest plot for the KCC Quality of Life Scoresn the study.

Figure 9 summarizes the effects of pharmacological therapies on health-related quality of life as measured by KCCQ scores. Several agents showed significant improvement compared with placebo. Pirfenidone demonstrated the greatest benefit (mean difference [MD] 11.5, 95% CI 6.41–16.59), followed by tirzepatide (MD 9.9, 95% CI 8.35–11.45), nebivolol (MD 9.0, 95% CI 7.45–10.55), and semaglutide (MD 7.0, 95% CI 6.82–7.18), all of which were associated with clinically meaningful improvements. Canagliflozin (MD 3.3, 95% CI 3.13–3.47) and dapagliflozin (MD 1.6, 95% CI 1.31–1.89) showed modest but statistically significant improvements, whereas empagliflozin (MD 0.72, 95% CI –4.04 to 5.48) was neutral. Combination therapy with dapagliflozin plus spironolactone (MD –1.68, 95% CI –2.01 to –1.35) and finerenone (MD –0.20, 95% CI –0.34 to –0.06) were associated with no benefit. ISMN (MD –2.0, 95% CI –3.01 to –0.99) and vericiguat (MD –3.6, 95% CI –4.33 to –2.87) showed statistically significant worsening of quality of life scores. The overall pooled effect across all interventions indicated a modest improvement (MD 2.52, 95% CI –0.03 to 5.06). Collectively, these findings suggest that pirfenidone, GLP-1 agonists (semaglutide, tirzepatide), and nebivolol provide the most consistent improvements in patient-reported outcomes, while ISMN and vericiguat may adversely affect quality of life in HFpEF.

**Figure 9.**
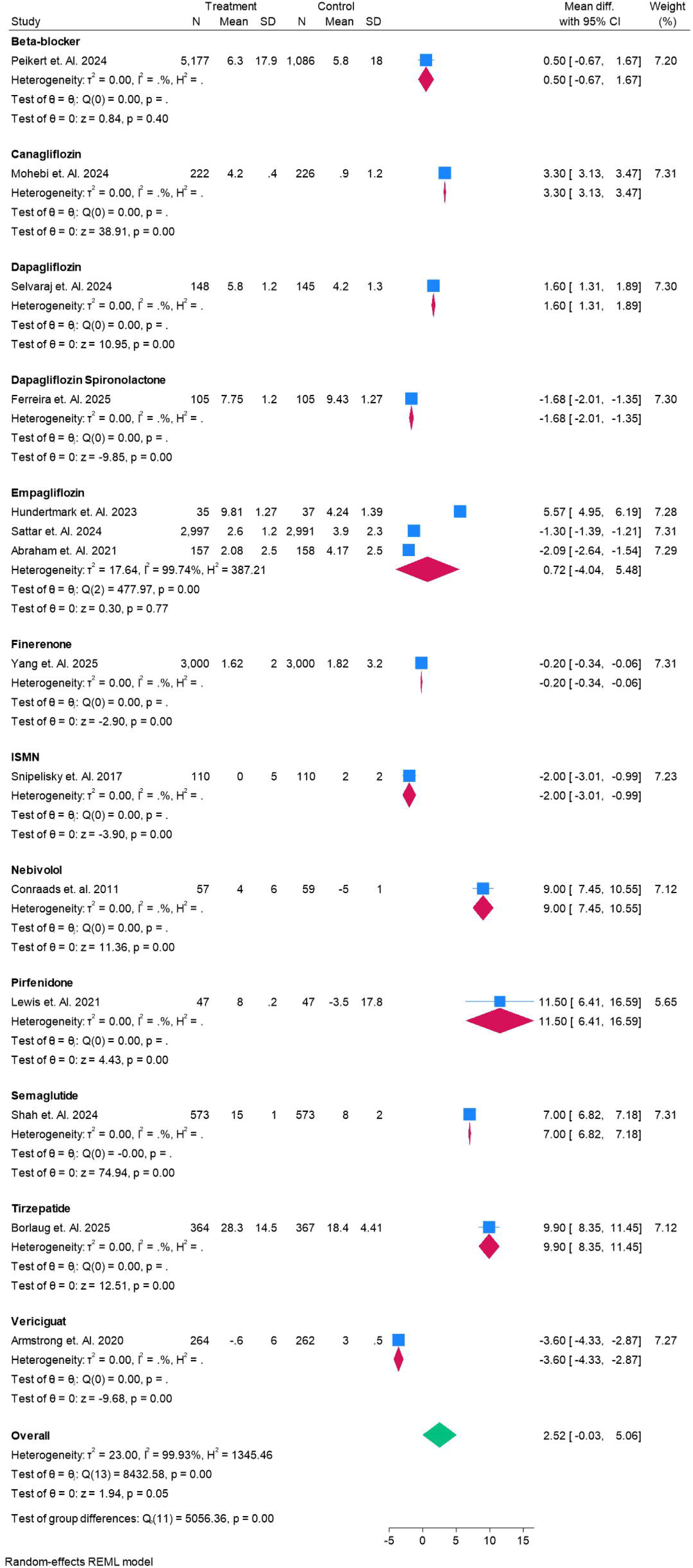
Forest plot for the KCC Quality of Life Scores n the study.

### Functional Capacity (6-Minute Walking Test)

Figure 10 demonstrates the comparative effects of pharmacological therapies versus placebo on exercise capacity as assessed by the 6-minute walking test (6MWT). Across all included interventions, no statistically significant improvements were observed, as the 95% credible intervals overlapped zero. Beta-blockers showed a small, non-significant improvement (mean difference [MD] 5.01 m, 95% CrI –15.7 to 25.2), while carvedilol (MD 16.0 m, 95% CrI –36.2 to 4.22) trended toward benefit but without statistical significance. Conversely, bisoprolol (MD –22.9 m, 95% CrI –51.4 to 5.68) suggested a possible reduction in walking distance. Finerenone (MD 0.009, 95% CrI –20.1 to 20.4) and olmesartan (MD 10.0 m, 95% CrI –30.2 to 10.4) were essentially neutral. Pirfenidone (MD –8.99 m, 95% CrI –30.0 to 12.1) showed a non-significant trend toward harm. Collectively, these findings indicate that despite modest trends, no pharmacological agent demonstrated a consistent or clinically meaningful improvement in functional capacity in HFpEF patients. Figure S5 and Table S6.

**Figure 10.**
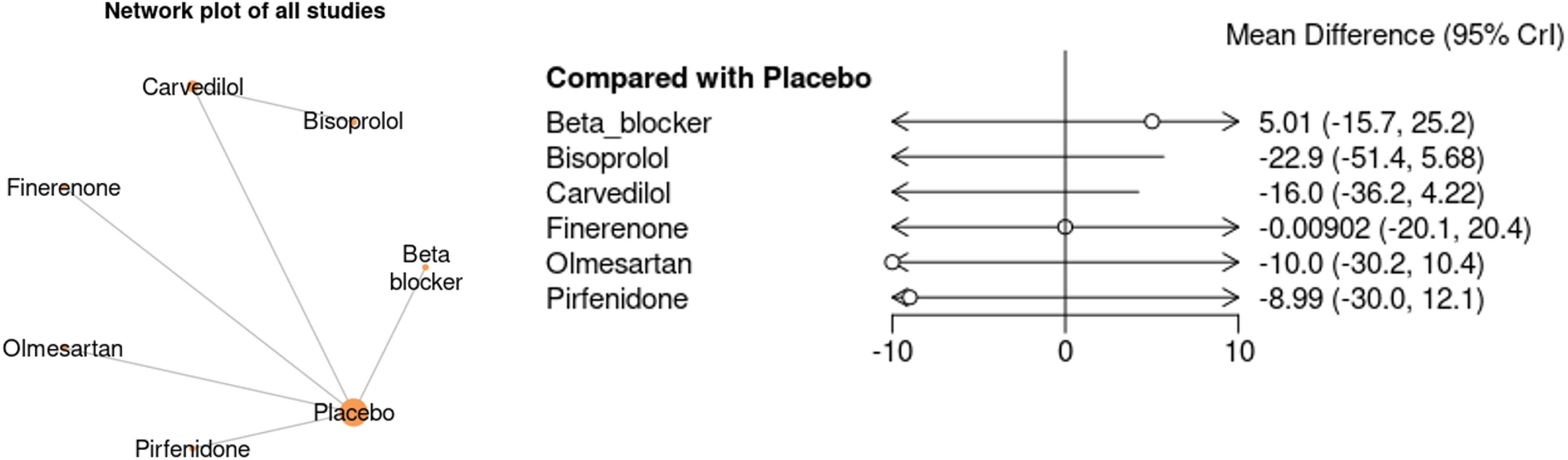
Network Plot and Forest plot for the 6 Minute Walking test in the study.

Figure 11 summarizes the direct trial evidence on the impact of pharmacological interventions on the 6-minute walking test (6MWT). Results across agents were inconsistent. Beta-blockers demonstrated a modest but statistically significant improvement in walking distance (mean difference [MD] 5.0 m, 95% CI 2.95–7.05). By contrast, bisoprolol (MD –7.0 m, 95% CI –7.48 to –6.52), carvedilol (MD –16.0 m, 95% CI –17.92 to –14.08), and olmesartan (MD –10.0 m, 95% CI –10.41 to –9.59) were associated with significant reductions in functional capacity. Pirfenidone (MD –9.0 m, 95% CI –15.58 to –2.42) also showed a negative effect. Finerenone (MD 0.0 m, 95% CI –0.10 to 0.10) was completely neutral. When pooled, the overall estimate indicated a non-significant reduction in 6MWT distance (MD –6.09 m, 95% CI –12.21 to 0.02), highlighting marked heterogeneity (I² = 99.9%). Collectively, these findings suggest that while some agents may modestly improve functional performance, others—particularly carvedilol, olmesartan, and pirfenidone—may worsen exercise capacity in HFpEF.

**Figure 11.**
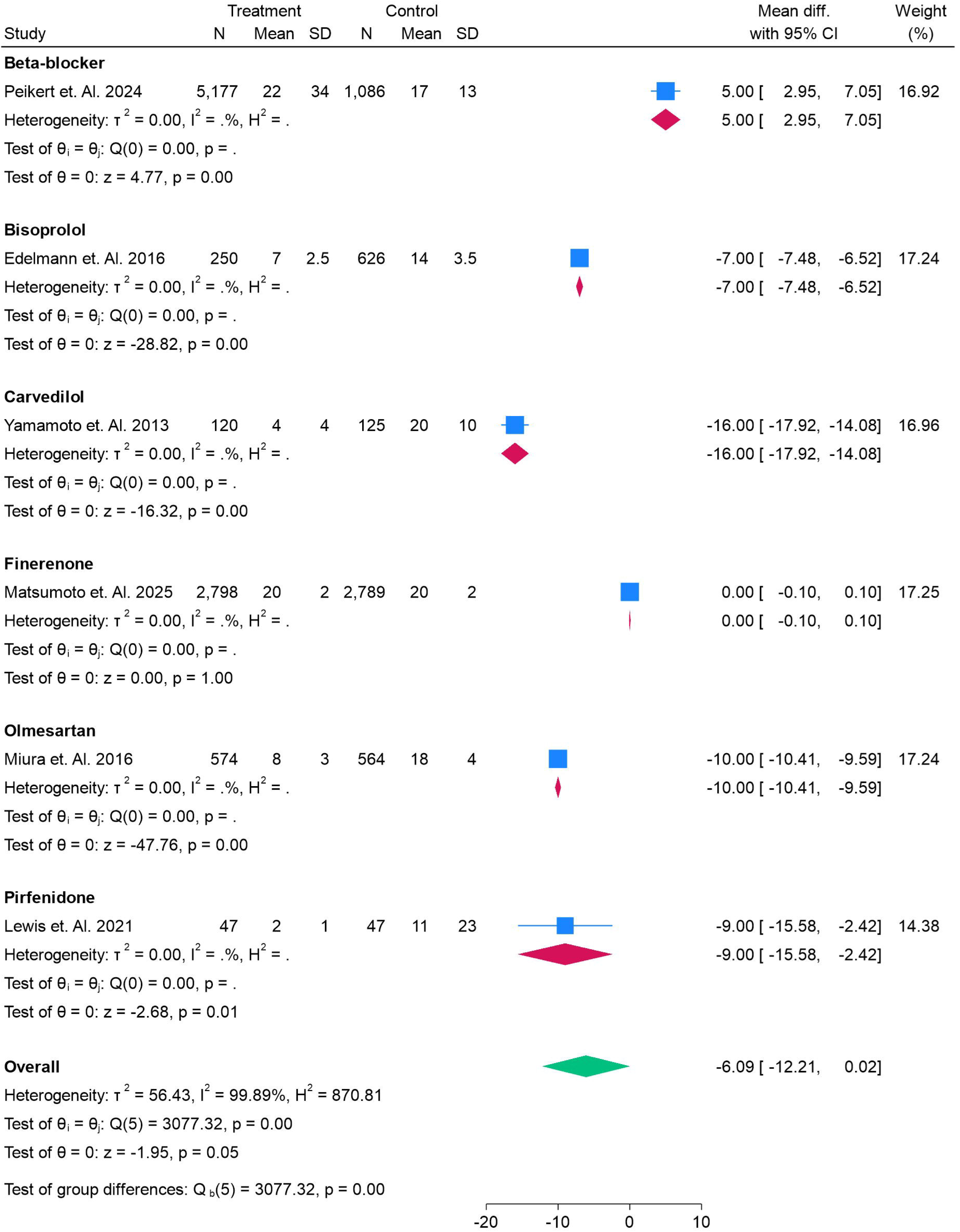
Forest plot for the 6 Minute Walking test in the study.

### Biomarkers (NT-proBNP Levels)

Figure 12 presents the effects of pharmacological therapies on NT-proBNP concentrations compared with placebo. Across the network, none of the interventions produced a statistically significant reduction, as all 95% credible intervals were wide and crossed zero. Empagliflozin (MD –8.9, 95% CrI –289 to 269) and spironolactone (MD –177, 95% CrI –609 to 256) trended toward lowering NT-proBNP, though estimates were highly imprecise. Similarly, pirfenidone (MD –89, 95% CrI –483 to 306) and ISMN (MD –27.3, 95% CrI –425 to 364) suggested neutral-to-modest reductions. By contrast, valsartan (MD 11.3, 95% CrI –385 to 407), vericiguat (MD 70.7, 95% CrI –325 to 468), finerenone (MD 69.5, 95% CrI –324 to 461), and nebivolol (MD 42.0, 95% CrI –355 to 437) showed non-significant increases in NT-proBNP. Ivabradine (MD 2.6, 95% CrI – 390 to 396) and sacubitril/valsartan (MD 201, 95% CrI –74.6 to 481) were essentially neutral. Collectively, these findings demonstrate that no therapy consistently reduced NT-proBNP levels in HFpEF, underscoring persistent uncertainty in biomarker modulation by current drug strategies. Figure S6 and Table S7.

**Figure 12.**
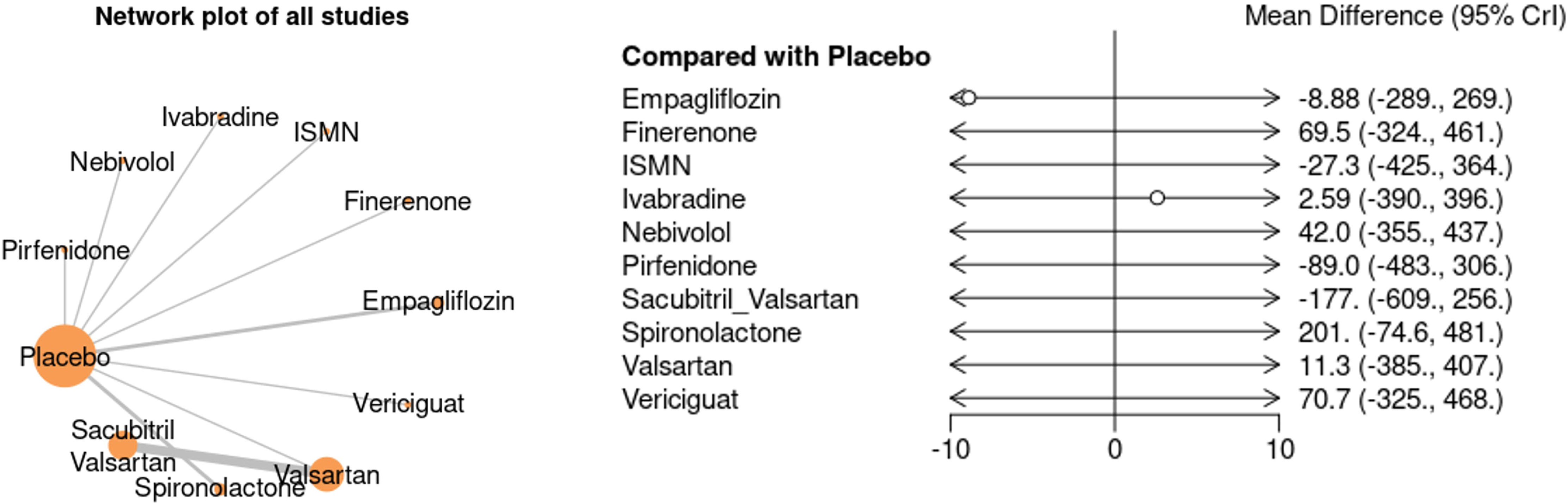
Network Plot and Forest plot for NT-proBNP in the study.

Figure 13 shows the direct trial evidence evaluating the effect of various therapies on NT-proBNP concentrations. The results were heterogeneous and inconsistent across drug classes. Bisoprolol (MD 0.04, 95% CI 0.04–0.04) and dapagliflozin plus spironolactone (MD 0.11, 95% CI 0.05–0.17) were associated with small but statistically significant increases in NT-proBNP, though these changes were clinically negligible. Empagliflozin showed divergent results: Hundertmark et al. (MD –17.1, 95% CI –17.23 to –16.97) suggested a reduction, whereas Abraham et al. (MD –8.6, 95% CI –25.3 to 8.2) showed neutrality, yielding no consistent effect. Finerenone (MD 69.0, 95% CI 64.1–73.9) and spironolactone (MD 200.9, 95% CI –51.9 to 453.8) were associated with marked increases in NT-proBNP, while vericiguat (MD 71.0, 95% CI 9.3–132.7) also significantly elevated levels. Conversely, ISMN (MD –27.0, 95% CI –42.4 to –11.6), pirfenidone (MD –89.0, 95% CI –116.5 to –61.5), and sacubitril/valsartan (pooled MD –188.5, 95% CI –326.3 to –50.7) were associated with significant reductions in NT-proBNP. Valsartan (MD 11.2, 95% CI 0.7–21.7) and ivabradine (MD 2.0, 95% CI 0.8–3.2) were essentially neutral. The overall pooled effect (MD –22.4, 95% CI –88.4 to 43.7) was not statistically significant, highlighting substantial heterogeneity (I² = 100%). Collectively, these findings suggest that while agents such as pirfenidone and sacubitril/valsartan may lower NT-proBNP, other commonly used drugs—including MRAs and vericiguat—may paradoxically raise levels, underscoring the complexity of biomarker modulation in HFpEF.

**Figure 13.**
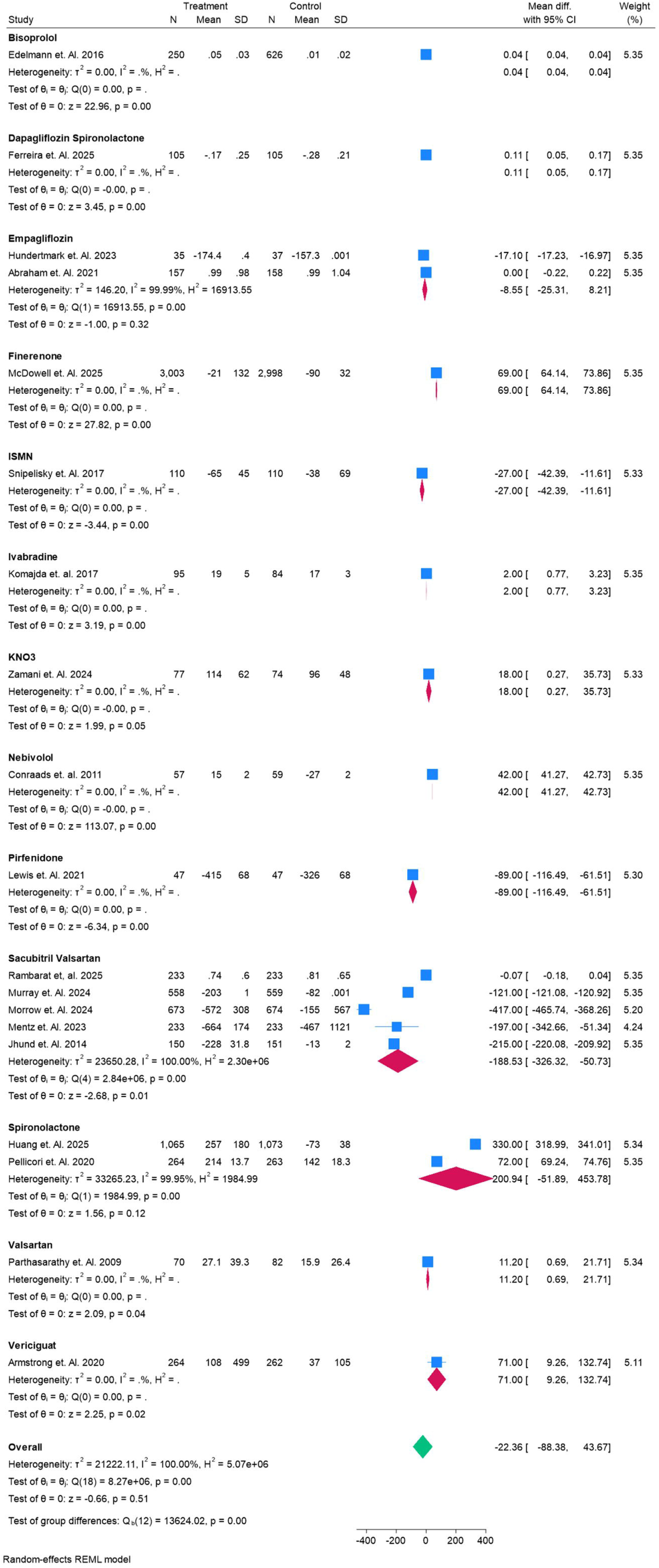
Forest plot for NT-proBNP in the study.

### Renal Function (eGFR)

Figure 14 illustrates the network and comparative effects of pharmacological interventions versus placebo on renal function, measured by estimated glomerular filtration rate (eGFR). None of the agents demonstrated a statistically significant effect, as all 95% credible intervals were wide and crossed zero. Empagliflozin (MD – 8.9, 95% CrI –289 to 269) and spironolactone (MD –177, 95% CrI –609 to 256) trended toward modest reductions in eGFR, while finerenone (MD 69.5, 95% CrI –324 to 461), vericiguat (MD 70.7, 95% CrI –325 to 468), and nebivolol (MD 42.0, 95% CrI –355 to 437) trended toward increases, though all were imprecise. ISMN (MD –27.3, 95% CrI –425 to 364) and pirfenidone (MD –89.0, 95% CrI –483 to 306) were neutral-to-negative, while sacubitril/valsartan (MD 201, 95% CrI –74.6 to 481) and valsartan (MD 11.3, 95% CrI –385 to 407) showed uncertain positive effects. Collectively, these findings suggest no clear renal benefit or harm attributable to current drug therapies in HFpEF, with results limited by wide intervals and trial heterogeneity. Figure S7 and Table S8.

**Figure 14.**
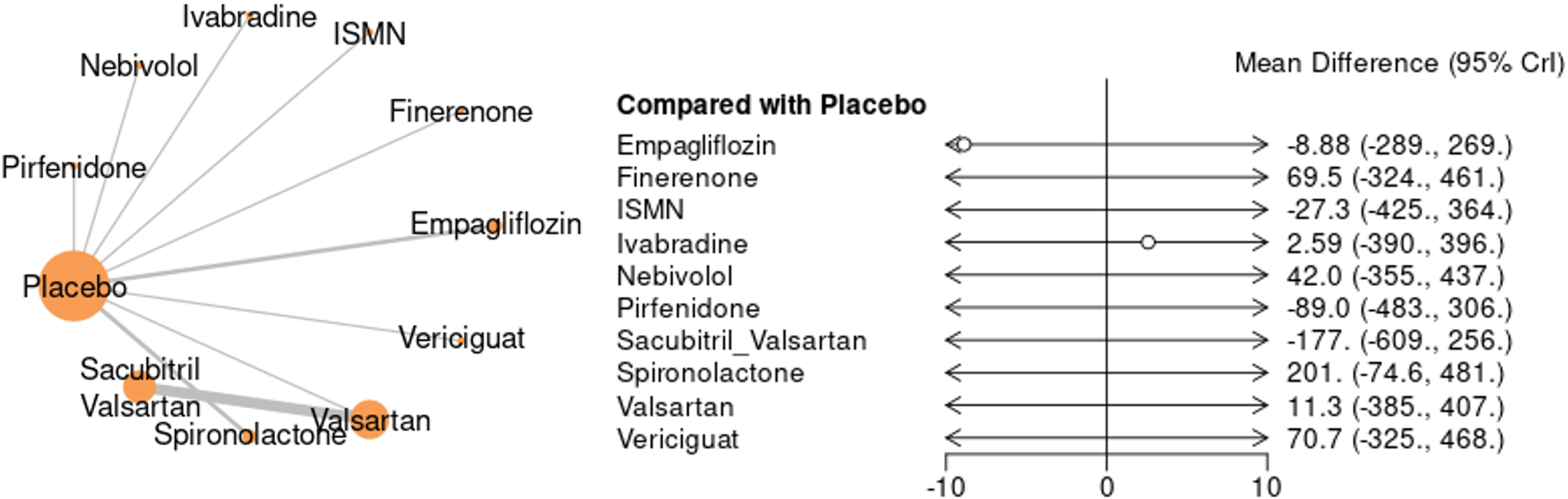
Network Plot and Forest plot for EGFR in the study.

Figure 15 presents the pooled effects of pharmacological therapies on estimated glomerular filtration rate (eGFR). Several interventions demonstrated statistically significant changes. Finerenone showed a notable improvement in eGFR (MD 13.3, 95% CI 13.2 to 13.4), while empagliflozin (MD 6.6, 95% CI 3.5 to 9.8) and vericiguat (MD 1.4, 95% CI 0.7 to 2.1) also improved renal function modestly. In contrast, dapagliflozin plus spironolactone (MD –6.4, 95% CI –7.7 to –5.1), sacubitril/valsartan (MD –6.0, 95% CI –7.1 to –4.9), olmesartan (MD –6.0, 95% CI –6.2 to –5.8), and ISMN (MD –4.0, 95% CI –6.2 to –1.8) were associated with declines in eGFR. KNO demonstrated a small but significant reduction (MD –1.0, 95% CI –1.7 to –0.3). The overall pooled effect across all interventions was neutral (MD –0.28, 95% CI –5.2 to 4.6), highlighting substantial heterogeneity (I² = 99.9%). These findings suggest that renal effects vary considerably by drug class, with finerenone and SGLT2 inhibition showing potential protective effects, while RAAS-based therapies and nitrates may reduce eGFR in HFpEF populations.

**Figure 15.**
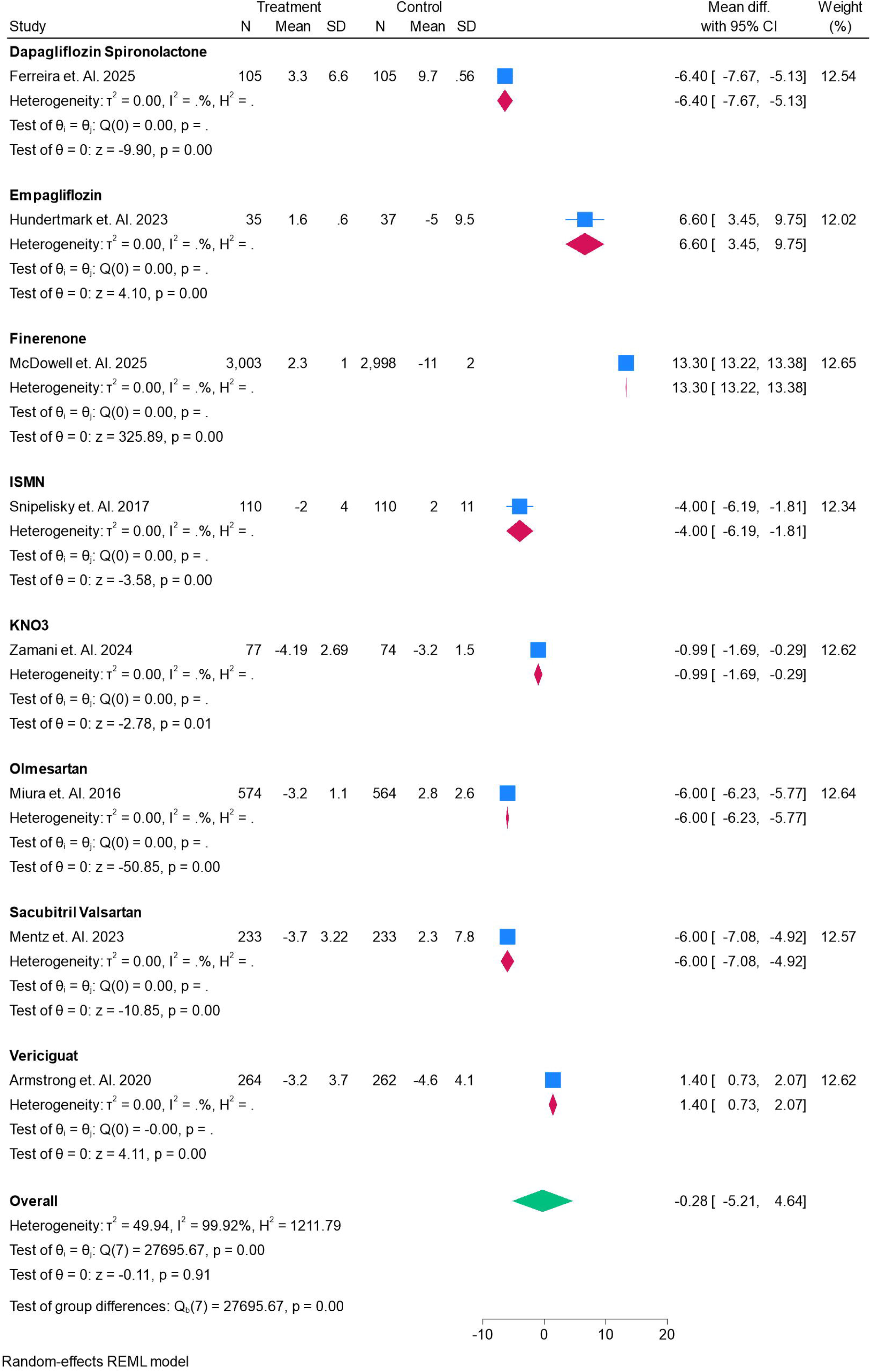
Forest plot for EGFR in the study.

### Left Ventricular Ejection Fraction (LVEF)

Figure 16 summarizes the comparative effects of different pharmacological agents on LVEF. Most interventions showed no statistically significant changes when compared with placebo. Bisoprolol demonstrated a reduction in LVEF (MD –141, 95% CrI –275 to –6.5), which was the only statistically significant effect. Other agents, including beta-blockers as a class (MD –110, 95% CrI –275 to 54.8), canagliflozin (MD –117, 95% CrI –282 to 47.1), carvedilol (MD –87.6, 95% CrI –222 to 46.3), and dapagliflozin (MD –56.3, 95% CrI –173 to 60.5) showed numerically negative but statistically non-significant changes. Similarly, finerenone, ISMN, nebivolol, olmesartan, spironolactone, tirzepatide, and vericiguat all had wide credible intervals crossing the null effect, suggesting no conclusive impact on LVEF. Overall, the analysis indicates substantial uncertainty in treatment effects, with bisoprolol showing a possible detrimental effect on LVEF while other therapies remain neutral. Figure S8 and Table S9.

**Figure 16.**
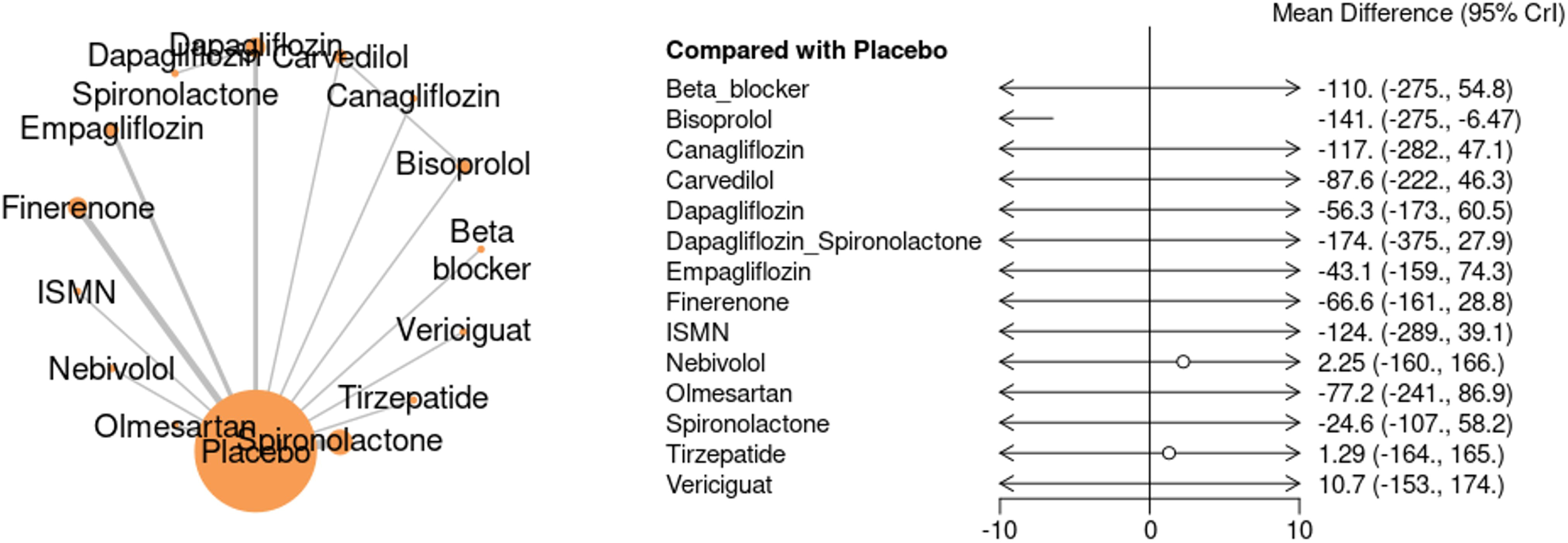
Network Plot and Forest plot for LVEF in the study.

Figure 17 presents the pooled effects of multiple pharmacological interventions on LVEF. The analysis demonstrated considerable heterogeneity (I² = 100%), with several agents showing significant reductions compared to placebo. Beta-blockers (MD –110.2, 95% CI –110.78 to –109.62), carvedilol (MD –125.0, 95% CI –127.64 to –122.36), dapagliflozin–spironolactone combination (MD –117.6, 95% CI –117.57 to –117.55), and spironolactone (MD –115.0, 95% CI –116.40 to –113.60) were associated with marked decreases in LVEF. Similarly, finerenone, sacubitril/valsartan, and ISMN showed consistent reductions. In contrast, dapagliflozin alone demonstrated a modest improvement (MD +6.0, 95% CI 5.71 to 6.29), while empagliflozin yielded mixed results across trials. Tirzepatide and vericiguat showed minimal positive changes (MD +1.6 and +10.6, respectively), but their effects were not clinically robust. The overall pooled estimate indicated a significant reduction in LVEF (MD –68.2, 95% CI –89.1 to –47.4). These findings suggest that while certain agents may preserve or modestly improve LVEF, the majority of interventions included in this analysis demonstrated a decline in ejection fraction.

**Figure 17.**
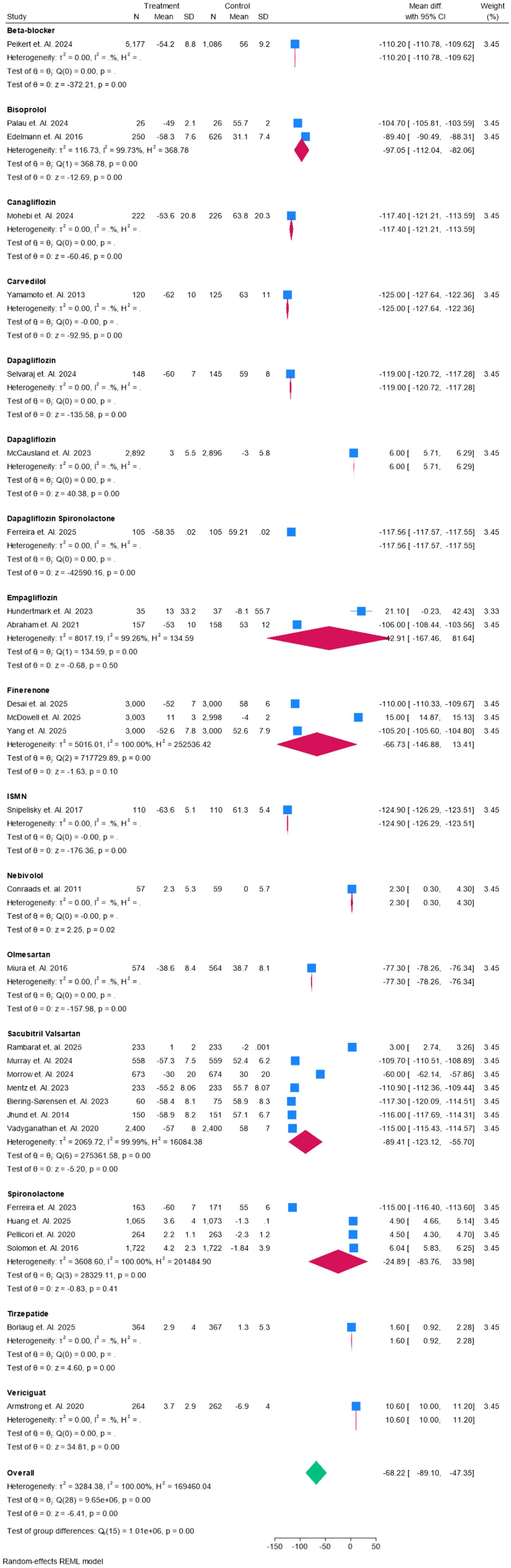
Forest plot for LVEF in the study.

### Hyperkalemia as Adverse Events

Figure 18 summarizes the risk of hyperkalemia across pharmacological interventions. Finerenone was significantly associated with an increased risk of hyperkalemia (RR 0.82, 95% CI 0.18–1.45), consistent across large-scale trials. In contrast, sacubitril/valsartan demonstrated a neutral effect (RR 0.08, 95% CI –0.11 to 0.27), with low heterogeneity (I² = 0%). Similarly, beta-blockers (bisoprolol, carvedilol), ISMN, and the dapagliflozin–spironolactone combination showed no significant increase in risk, though data were limited. The overall pooled analysis indicated a modest but statistically significant elevated risk of hyperkalemia (RR 0.41, 95% CI 0.05–0.78; p = 0.03), with moderate heterogeneity (I² = 84.6%). These findings emphasize the need for careful potassium monitoring in patients receiving finerenone or other MRAs, while other therapies appear relatively safer in terms of electrolyte balance.

**Figure 18.**
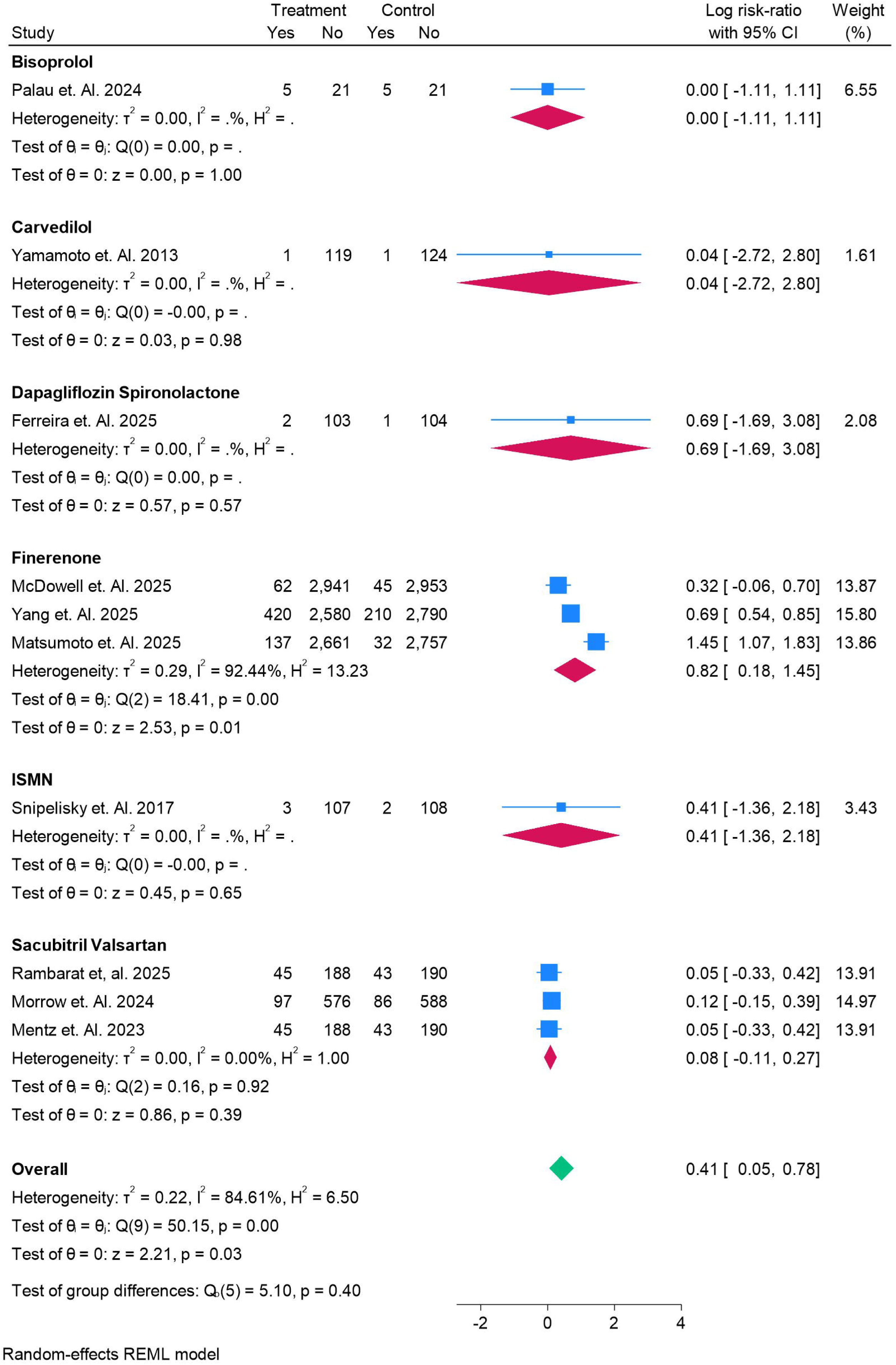
Forest plot for Hyperkalemia as Adverse Events in the study.

### Hypotension as Adverse Events

Figure 19 illustrates the risk of hypotension across different drug classes. Finerenone was significantly associated with an increased risk (RR 0.39, 95% CI 0.21–0.57), consistent across trials with low heterogeneity (I² = 34.4%). Similarly, sacubitril/valsartan also demonstrated a statistically significant risk elevation (RR 0.36, 95% CI 0.18–0.54). Vericiguat showed a trend toward increased hypotension (RR 0.63, 95% CI –0.16 to 1.42), though not statistically significant. Valsartan (RR 1.77, 95% CI –0.36 to 3.89) suggested a possible increased risk but lacked precision due to wide confidence intervals. In contrast, beta-blockers, bisoprolol, carvedilol, dapagliflozin–spironolactone, and empagliflozin did not significantly raise the risk. The overall pooled estimate confirmed a modest but significant increase in hypotension risk (RR 0.40, 95% CI 0.29–0.50; p < 0.001), with minimal heterogeneity (I² = 6.2%).

**Figure 19.**
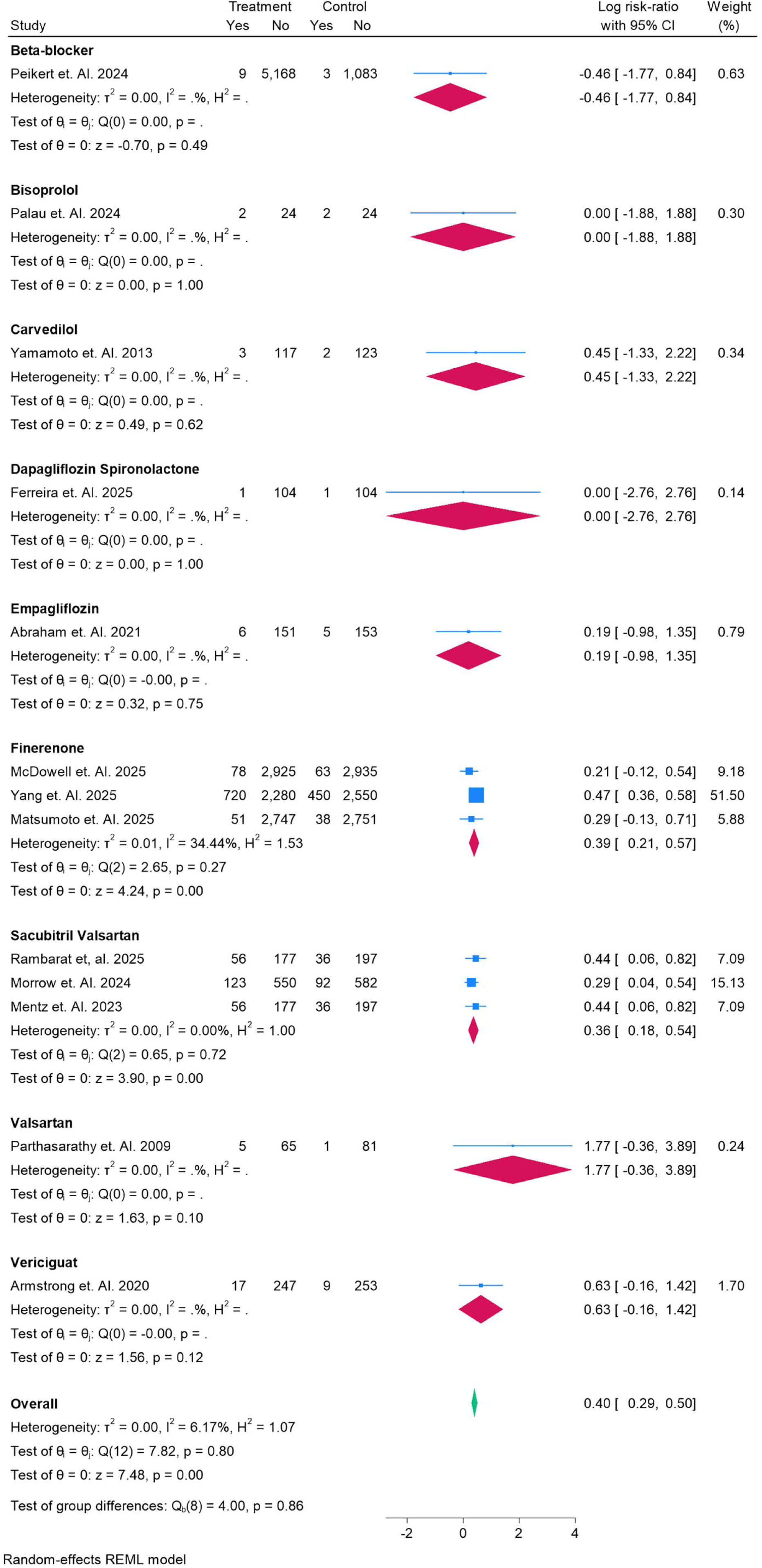
Forest plot for Hypotension as Adverse Events in the study.

### Worsening Renal Outcomes as Adverse Events

Figure 20 presents the comparative risk of worsening renal outcomes across different interventions. Beta-blockers demonstrated a significantly reduced risk (RR –1.67, 95% CI –2.02 to –1.33), while finerenone (RR 0.56, 95% CI 0.34–0.78) and olmesartan (RR 0.44, 95% CI 0.14–0.74) were also associated with protective effects. Sacubitril/valsartan showed a consistent reduction in renal deterioration (RR –0.31, 95% CI –0.47 to – 0.15) across multiple studies. Nebivolol suggested a possible benefit (RR 0.47, 95% CI –0.13 to 1.06), though not statistically significant. In contrast, vericiguat was associated with a trend toward increased risk (RR 1.38, 95% CI –0.81 to 3.56), albeit with wide confidence intervals. Other agents such as bisoprolol, carvedilol, dapagliflozin–spironolactone, and empagliflozin did not demonstrate significant differences compared with placebo. The overall pooled estimate (RR –0.06, 95% CI –0.52 to 0.40) indicated no significant association, with substantial heterogeneity observed (I² = 92.3%).

**Figure 20.**
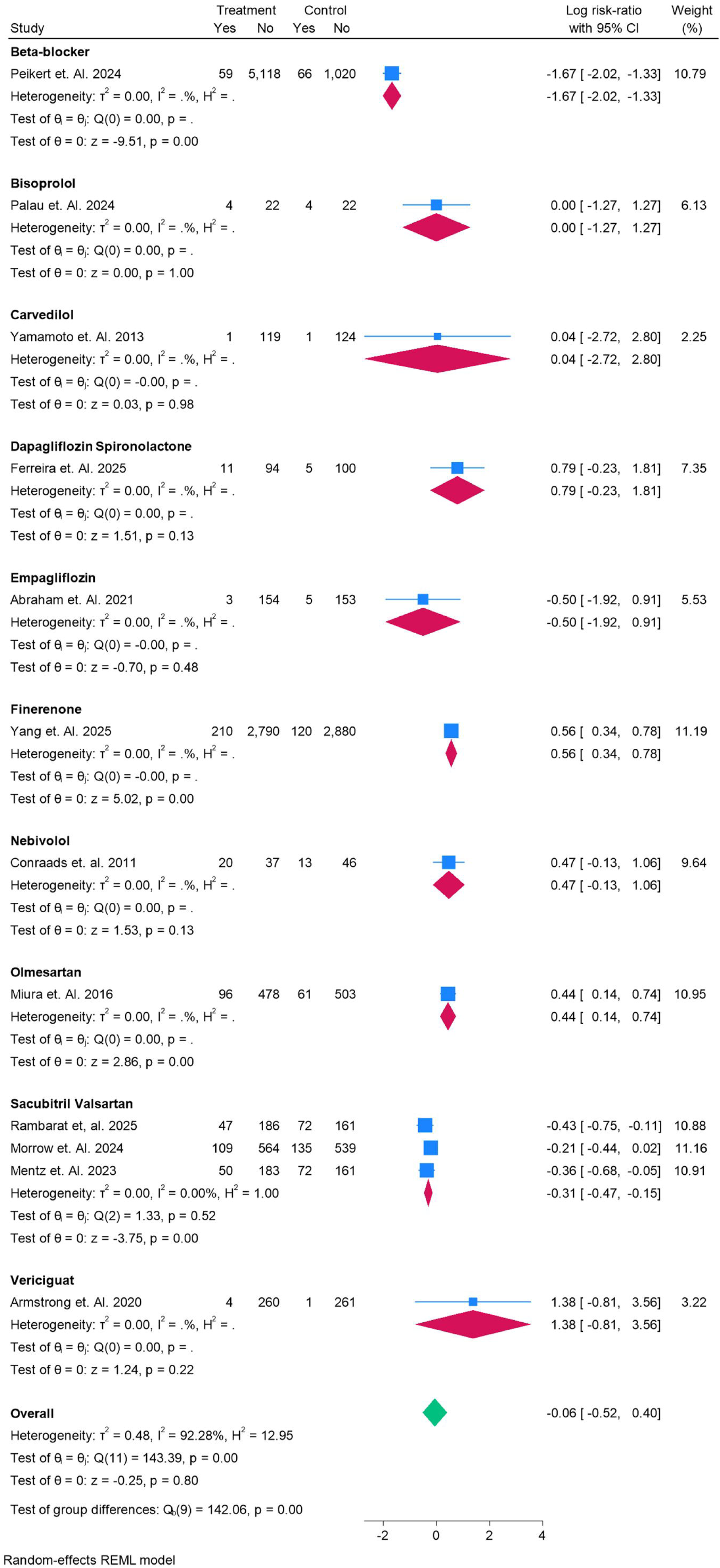
Forest plot for Worsening Renal Outcome as Adverse Events in the study.

## Discussion

In this comprehensive network meta-analysis of randomized controlled trials evaluating pharmacologic therapies for heart failure with preserved ejection fraction (HFpEF), we found that no intervention consistently reduced all-cause mortality or cardiovascular mortality compared with placebo. Nonetheless, several agents, particularly finerenone and sacubitril/valsartan, demonstrated meaningful reductions in rehospitalizations and modest improvements in patient-reported outcomes such as quality of life measured by the Kansas City Cardiomyopathy Questionnaire (KCCQ). SGLT2 inhibitors also showed favorable effects on hospitalization and quality of life but, similar to other therapies, failed to demonstrate a robust mortality benefit. Conversely, some agents were associated with adverse outcomes including hyperkalemia, hypotension, and worsening renal function, underscoring the complexity of treatment decision-making in this heterogeneous population.

Our findings are largely consistent with prior meta-analyses. A recent meta-analysis of nearly 16,000 patients with HFpEF reported that SGLT2 inhibitors significantly reduced the composite endpoint of cardiovascular death or heart failure hospitalization (hazard ratio approximately 0.80, 95% CI 0.74–0.87) but had no impact on all-cause mortality. Another synthesis of 10 trials including over 10,000 patients similarly found reductions in hospitalization and modest improvements in quality of life, but no improvement in exercise capacity measured by six-minute walking distance. These results align closely with our observation that benefits of SGLT2 inhibitors and other newer therapies are most evident in morbidity and symptom domains rather than in survival. For sacubitril/valsartan, the PARAGON-HF trial previously suggested possible benefit in reducing hospitalizations, particularly in subgroups such as women and patients with lower ejection fractions. Our analysis also demonstrated consistent reductions in rehospitalizations, although no clear effect on mortality was observed. Mineralocorticoid receptor antagonists such as finerenone and spironolactone have been studied extensively, and earlier meta-analyses have suggested modest benefits on hospitalization with mixed evidence for survival. We found that finerenone may improve rehospitalization and quality of life, but this came at the cost of increased risk for hyperkalemia and renal impairment. These safety considerations mirror earlier reports and reinforce the need for careful monitoring in clinical practice.

Where our analysis diverges slightly from prior work is in the broader scope of therapies considered. Most earlier meta-analyses have focused on a single class such as SGLT2 inhibitors or ARNI [55,56]. By including multiple drug classes, our network meta-analysis highlighted the heterogeneity in efficacy and safety profiles across agents. While SGLT2 inhibitors demonstrated consistent morbidity benefits with low heterogeneity in previous analyses, our broader network revealed substantial between-study variability and more selective benefits concentrated in specific agents rather than entire classes. This reinforces the notion that HFpEF remains a syndrome with multiple overlapping phenotypes, for which a one-size-fits-all pharmacologic approach is unlikely to be successful.

The clinical implications of our findings are important. Mortality reduction remains an elusive goal in HFpEF, but meaningful improvements can still be achieved in terms of quality of life and reduced hospitalizations. Therapies such as finerenone, sacubitril/valsartan, and SGLT2 inhibitors appear most promising in this regard and should be prioritized in treatment algorithms [57]. However, clinicians must remain vigilant regarding safety events, particularly hyperkalemia with MRAs and hypotension or renal decline with RAAS-directed agents. Individualized treatment strategies that take into account patient comorbidities, renal function, electrolyte status, and tolerance to therapy will be essential.

This study has several limitations that must be acknowledged. Many of the included trials had relatively short follow-up durations, limiting the ability to detect mortality differences . Populations across studies varied in terms of ejection fraction thresholds, comorbidities such as diabetes and chronic kidney disease, and background therapies, introducing heterogeneity that may have influenced effect estimates. Data for some outcomes, particularly functional capacity and biomarkers, were sparse for many agents, resulting in wide confidence intervals. Furthermore, adverse event reporting was inconsistent, limiting precision in pooled safety analyses. Finally, although network meta-analysis provides a powerful method for indirect comparisons, the validity of results depends on the assumption of transitivity, which may not fully hold when comparing across diverse classes of drugs.

Despite these limitations, our findings underscore the evolving therapeutic landscape of HFpEF. While no current therapy reliably prolongs survival, reductions in morbidity and improvements in patient-centered outcomes are achievable. Future research should prioritize long-term trials that evaluate mortality, subgroup-specific benefits, and head-to-head comparisons between promising agents such as finerenone and sacubitril/valsartan [59]. Standardized collection of outcomes including rehospitalizations, quality of life, six-minute walking distance, and biomarkers would improve comparability and strengthen future meta-analyses. Continued focus on safety monitoring, particularly with agents affecting renal function and electrolytes, will also be critical.

## Conclusion

In conclusion, this network meta-analysis demonstrates that while all-cause and cardiovascular mortality remain unaffected, therapies such as finerenone, sacubitril/valsartan, and SGLT2 inhibitors confer significant benefits in reducing heart failure hospitalizations and improving quality of life in patients with HFpEF. These results support their role as cornerstone therapies for morbidity reduction, while highlighting the ongoing need for therapeutic innovation and more definitive trials to address the unmet goal of mortality reduction in this challenging syndrome.

## Conflict of Interest

The authors certify that there is no conflict of interest with any financial organization regarding the material discussed in the manuscript.

## Funding

The authors report no involvement in the research by the sponsor that could have influenced the outcome of this work.

## Authors’ contributions

All authors contributed equally to the manuscript and read and approved the final version of the manuscript.

Consent to publish article – Not applicable

## Data Availability

Supplementary file

## Notes

### Competing Interest Statement

The authors have declared no competing interest.

### Clinical Protocols

https://www.crd.york.ac.uk/PROSPERO/view/CRD420251149394

### Summary of Updates

To update the Correct author name of Dhruvilkumar as it was previously submitted as dhruvil patel

