## supplementary file for "Efficacy and Safety of Pharmacologic Therapies in HFpEF: A Systematic Review and Network Meta-Analysis"

Supplementary Files

Search String

("Heart Failure with Preserved Ejection Fraction"[Mesh] OR HFpEF OR "diastolic heart failure" OR "preserved ejection fraction")

AND

(randomized controlled trial[pt] OR randomized[tiab] OR placebo[tiab] OR trial[ti])

AND

(

"Sodium-Glucose Transporter 2 Inhibitors"[Mesh] OR dapagliflozin OR empagliflozin OR sotagliflozin OR SGLT2i

OR "Angiotensin Receptor-Neprilysin Inhibitors"[Mesh] OR sacubitril OR valsartan OR ARNI

OR "Mineralocorticoid Receptor Antagonists"[Mesh] OR spironolactone OR eplerenone OR finerenone OR MRA

OR "Angiotensin-Converting Enzyme Inhibitors"[Mesh] OR perindopril OR enalapril OR lisinopril OR ACE inhibitor

OR "Angiotensin Receptor Antagonists"[Mesh] OR candesartan OR irbesartan OR losartan OR ARB

OR "Adrenergic beta-Antagonists"[Mesh] OR beta-blocker OR metoprolol OR bisoprolol OR carvedilol

OR diuretics OR furosemide OR torsemide OR bumetanide OR chlorthalidone

OR pirfenidone

OR vericiguat

OR ivabradine

OR digoxin

OR nitrates OR "isosorbide mononitrate"

OR sildenafil OR "phosphodiesterase 5 inhibitor"

)

Table S1. Summary Findings and baseline

| **Ref Num** | **Author Year** | **Country** | **Total Sample** | **Total Male** | **Total Female** | **Age** | **Follow Up** | **6 Minute Walk test in Treatment Group Mean** | **SD** | **6 Minute Walk test in ControlGroup Mean** | **Mean** | **Treatment Group EF** | **EF SD** | **Control Group EF** | **EFSD** | **Treatment Name** | **Treatment Number** | **Control Name** | **Control Number** | **Grade** | **Main Finding** |
| --- | --- | --- | --- | --- | --- | --- | --- | --- | --- | --- | --- | --- | --- | --- | --- | --- | --- | --- | --- | --- | --- |
|  | Rambarat et, al. 2025 | USA | 466 | 224 | 242 | 70 | 20 | N/A | N/A | N/A | N/A | 55 | 6 | 54 | 8 | Sacubitril Valsartan | 233 | Valsartan | 233 | High | Sacubitril/valsartan reduced NT-proBNP more than valsartan, with consistent efficacy and safety across men and women |
|  | Ferreira et. Al. 2023 | Multicenter | 525 | 390 | 135 | 73 | 9 | N/A | N/A | N/A | N/A | 60 | 7 | 55 | 6 | Spironolactone | 163 | Placebo | 171 | High | In patients with stage B HFpEF, spironolactone for 9 months shortened QRS duration, suggesting a beneficial effect on myocardial electrical activation independent of structural changes. |
|  | Parthasarathy et. Al. 2009 | Multicenter | 152 | 76 | 76 | 61 | 3.5 | 68 |  | 82 |  |  |  |  |  | Valsartan | 70 | Placebo | 82 | High | This multicenter RCT (n=152, UK/Germany/Austria/Switzerland, 2009) found that valsartan 320 mg daily in HFpEF patients did **not** improve exercise capacity or 6MWT distance versus placebo. LVEF, BNP, and QoL scores showed no significant differences. Valsartan reduced exercise systolic BP and Borg perceived exertion scores, and was generally well-tolerated, though hypotension was slightly more frequent. |
|  | Xing et. Al. 2025 | China | 609 | 293 | 316 | 74 | 3 | N/A | N/A | N/A | N/A |  |  |  |  | Spironolactone | 304 | Placebo | 305 | High | Patients with **recurrent deterioration depression trajectories** had a **significantly greater risk of all-cause mortality** compared to those with low trajectories |
|  | Desai et. al. 2025 | Multicenter | 6001 | 2490 | 3511 | 76 | 32 | N/A | N/A | N/A | N/A | 52 | 7 | 58 | 6 | Finerenone | 3000 | Placebo | 3000 | High | Among patients with HFmrEF/HFpEF, about half of deaths were cardiovascular (mainly sudden death or HF progression), and **finerenone did not significantly reduce cause-specific mortality compared with placeb** |
|  | Palau et. Al. 2024 | Spain | 52 | 21 | 31 | 73 | 1 | N/A | N/A | N/A | N/A | 63 | 7.1 | 65.7 | 7 | Bisoprolol | 26 | Placebo | 26 | High | In patients with HFpEF and chronotropic incompetence, **β-blocker withdrawal improved functional capacity (peak VO₂), especially in those with smaller left ventricular systolic volumes** |
|  | McDowell et. Al. 2025 | Multicenter | 6001 | 3269 | 2732 | 72 | 3 | N/A | N/A | N/A | N/A | 52 | 7 | 58 | 6 | Finerenone | 3003 | Placebo | 2998 | High | **Finerenone reduced cardiovascular death and HF hospitalization consistently across all baseline risk groups in HFpEF/HFmrEF, with greatest absolute benefit in high-risk patients** |
|  | Armstrong et. Al. 2020 | Multicenter | 789 | 404 | 385 | 72.7 | 6 | 230 |  | 212 |  | 58.6 | 7.9 | 56.3 | 7 | Vericiguat | 264 | Placebo | 262 | High | Vericiguat did **not improve quality of life (KCCQ PLS) or 6MWT** in HFpEF after 24 weeks compared with placebo |
|  | Zamani et. Al. 2024 | USA | 84 | 26 | 58 | 68 | 1.2 | 63 |  | 52 |  | 61.6 | 0.5 | 60.9 | 0.5 | KNO3 | 77 | KCL | 74 | Moderate | Chronic potassium nitrate supplementation did **not improve exercise capacity, quality of life, or vasodilatory reserve** in HFpEF |
|  | Ferreira et. Al. 2025 | Portugal | 108 | 46 | 62 | 76 | 4 | N/A | N/A | N/A | N/A | 59 | 0.6 | 60.2 | 0.6 | Dapagliflozin Spironolactone | 105 | Dapagliflozin | 105 | High | Adding spironolactone to dapagliflozin in HFpEF/HFmrEF reduces NT-proBNP more effectively but increases risks of eGFR decline and hyperkalemia |
|  | Borlaug et. Al. 2025 | Multicenter | 731 | 402 | 329 | 62.9 | 12 | N/A | N/A | N/A | N/A | 60.4 | 6.5 | 61.5 | 6.5 | Tirzepatide | 364 | Placebo | 367 | High | Tirzepatide improved symptoms, exercise capacity, and reduced risk of HF events in obese HFpEF patients regardless of BMI/WHR, with greater benefit in those achieving larger weight loss. |
|  | Huang et. Al. 2025 | China | 2138 | 982 | 1156 | 67 | 40.8 | N/A | N/A | N/A | N/A | 56 | 6.5 | 57 | 6.2 | Spironolactone | 1065 | Placebo | 1073 | High | **Obesity and abdominal obesity increase the incidence of atrial fibrillation in HFpEF patients** |
|  | Miura et. Al. 2016 | Japan | 1138 | 851 | 287 | 64.7 | 53 | N/A | N/A | N/A | N/A | 38.6 | 8.4 | 38.7 | 8.1 | Olmesartan | 574 | Placebo | 564 | High | Olmesartan + β-blocker reduced mortality in hypertensive HFpEF patients, but triple therapy (Olmesartan + ACEI + BB) worsened outcomes in HFrEF |
|  | Conraads et. al. 2011 | Multicenter | 116 | 41 | 75 | 66 | 6 | 420 | 143 | 412 | 123 | 61.9 | 7.8 | 63.2 | 9.2 | Nebivolol | 57 | Placebo | 59 | High | Nebivolol did **not improve exercise capacity, quality of life, or NT-proBNP levels compared with placebo** in HFpEF patients, despite reductions in heart rate and blood pressure |
|  | Pellicori et. Al. 2020 | Multicenter | 527 | 392 | 135 | 73 | 9 | N/A | N/A | N/A | N/A | 63 | 5.3 | 63 | 5.3 | Spironolactone | 264 | Placebo | 263 | High | The HOMAGE trial enrolled 527 older patients at high risk of heart failure, showing they had preserved LVEF but elevated NT-proBNP and fibrosis markers, setting the stage to test whether spironolactone can reduce fibrotic remodeling |
|  | Solomon et. Al. 2016 | Multicenter | 3444 | 1669 | 1775 | 68.6 | 41 | N/A | N/A | N/A | N/A | 57.1 | 5.2 | 58.3 | 5.1 | Spironolactone | 1722 | Placebo | 1722 | High | In HFpEF patients from TOPCAT, spironolactone did not improve the composite primary outcome overall, but showed potential benefit at lower LVEF ranges, particularly for reducing HF hospitalizations |
|  | Yamamoto et. Al. 2013 | Japan | 245 | 142 | 103 | 72 | 38 | N/A | N/A | N/A | N/A | 62 | 10 | 63 | 11 | Carvedilol | 120 | Placebo | 125 | High | Carvedilol did not significantly improve outcomes in HFpEF overall, but standard-dose treatment suggested possible benefit |
|  | Edelmann et. Al. 2016 | Multicenter | 876 | 547 | 329 | 72.8 | 3 | N/A | N/A | N/A | N/A | 58.7 | 8.8 | 34.9 | 8 | Bisoprolol | 250 | Carvedilol | 626 | High | **Beta-blocker titration in elderly HFpEF patients was feasible but less tolerated, with more side effects and no significant functional or echocardiographic benefit compared to HFrEF** |
|  | Snipelisky et. Al. 2017 | USA | 110 | 40 | 59 | 69 | 3 | N/A | N/A | N/A | N/A | 63.6 | 5.1 | 61.3 | 5.4 | ISMN | 110 | Placebo | 110 | High | In HFpEF, **ISMN decreased daily physical activity without improving standard HF outcomes** |
|  | Murray et. Al. 2024 | Multicenter | 1117 | 539 | 578 | 73.5 | 32.4 | N/A | N/A | N/A | N/A | 57.3 | 7.5 | 52.4 | 6.2 | Sacubitril Valsartan | 558 | Valsartan | 559 | High | Numerous serum proteins (e.g., B2M, TIMP1, SVEP1, SERPINA4) were strongly associated with risk of HF hospitalization and CV death in HFpEF, but a proteomic risk score did not outperform established biomarkers (NT-proBNP, troponin). |
|  | Morrow et. Al. 2024 | USA | 1347 | 859 | 488 | 66 | 6 | N/A | N/A | N/A | N/A | 30 | 20 | 30 | 20 | Sacubitril Valsartan | 673 | Valsartan | 674 | Moderate | Sacubitril/valsartan reduced NT-proBNP levels and lowered the risk of cardiovascular death or hospitalization for HF compared with enalapril/valsartan in patients stabilized after worsening HF, particularly when LVEF ≤60% |
|  | Mentz et. Al. 2023 | USA | 466 | 224 | 242 | 70 | 7.9 | N/A | N/A | N/A | N/A | 55.2 | 8.06 | 55.7 | 8.07 | Sacubitril Valsartan | 233 | Valsartan | 233 | High | Sacubitril/valsartan produced a greater reduction in NT-proBNP than valsartan in HFpEF/HFmrEF with recent worsening HF, with more hypotension but fewer renal events |
|  | Biering-Sørensen et. Al. 2023 | Multicenter | 301 | 178 | 123 | 69.9 | 9 | N/A | N/A | N/A | N/A | 58.4 | 8.1 | 58.9 | 8.3 | Sacubitril Valsartan | 60 | Valsartan | 75 | Moderate | Sacubitril/valsartan improved global circumferential strain (GCS), but not global longitudinal strain (GLS), compared with valsartan over 36 weeks in HFpEF patients |
|  | Yang et. Al. 2025 | Multicenter | 6001 | 3277 | 2724 | 71.8 | 32 | N/A | N/A | N/A | N/A | 52.6 | 7.8 | 52.6 | 7.9 | Finerenone | 3000 | Placebo | 3000 | Moderate | **Finerenone reduced HF hospitalizations and CV events, and improved KCCQ quality-of-life scores, without increasing mortality** in HFmrEF/HFpEF. |
|  | Hundertmark et. Al. 2023 | Multicenter | 72 | 42 | 30 | 68.33 | 3 | N/A | N/A | N/A | N/A | 40 | 36 | 50 | 59 | Empagliflozin | 35 | Placebo | 37 | Moderate | Empagliflozin did **not improve myocardial energetics or circulating metabolites** after 12 weeks in HFrEF or HFpEF |
|  | McCausland et. Al. 2023 | Multicenter | 5788 | 3253 | 2535 | 72 | 36 | N/A | N/A | N/A | N/A | 55 | 9 | 54 | 9 | Dapagliflozin | 2892 | Placebo | 2896 | High | *An initial decline in eGFR with dapagliflozin is frequent but not associated with adverse cardiovascular or kidney outcomes, supporting continuation of therapy despite early eGFR dips.* |
|  | Mohebi et. Al. 2024 | USA | 448 | 247 | 201 | 65.4 | 3 | N/A | N/A | N/A | N/A | 53.6 | 20.8 | 63.8 | 20.3 | Canagliflozin | 222 | Placebo | 226 | High | *Canagliflozin rapidly improved health status (KCCQ-TSS) in both HFpEF and HFrEF, with the greatest benefit occurring in the first weeks after initiation* |
|  | Sattar et. Al. 2024 | Multicenter | 5998 | 3280 | 2700 | 73.5 | 26 | N/A | N/A | N/A | N/A |  |  |  |  | Empagliflozin | 2997 | Placebo | 2991 | Moderate | *Empagliflozin improved HF outcomes consistently across BMI categories in HFpEF, with no BMI-specific interaction* |
|  | Selvaraj et. Al. 2024 | Multicenter | 324 | 136 | 188 | 70 | 3 | 244 | 169 | 245 | 168 | 60 | 7 | 59 | 8 | Dapagliflozin | 148 | Placebo | 145 | Moderate | *Dapagliflozin improved HF symptoms and exercise capacity in HFpEF, but did not significantly alter systemic metabolomic pathways compared with placebo.* |
|  | Abraham et. Al. 2021 | Multicenter | 315 | 179 | 136 | 73.5 | 3 | 306 | 40 | 297 | 52 | 53 | 10 | 53 | 12 | Empagliflozin | 157 | Placebo | 158 | Moderate | Empagliflozin did not significantly improve 6MWT distance over 12 weeks in HF with reduced or preserved EF, but exploratory analyses suggested possible improvements in symptoms and diuretic use in HFrEF. |
|  | Jhund et. Al. 2014 | Multicenter | 301 | 129 | 172 | 71 | 9 | N/A | N/A | N/A | N/A | 58.9 | 8.2 | 57.1 | 6.7 | Sacubitril Valsartan | 150 | Valsartan | 151 | Moderate | LCZ696 reduced NT-proBNP, improved NYHA class, reduced LA size, and preserved renal function vs. valsartan in HFpEF, independently of blood pressure lowering |
|  | Vadyganathan et. Al. 2020 | Multicenter | 4796 | 2248 | 2548 | 72.7 | 35 | N/A | N/A | N/A | N/A | 57 | 8 | 58 | 7 | Sacubitril Valsartan | 2400 | Valsartan | 2400 | Moderate | Sacubitril/valsartan provided greater reduction in HF hospitalizations and CV death in HFpEF patients recently hospitalized compared to valsartan alone |
|  | Matsumoto et. Al. 2025 | Multicenter | 6001 | 3077 | 2510 | 71.7 | 32 | N/A | N/A | N/A | N/A |  |  |  |  | Finerenone | 2798 | Placebo | 2789 | Moderate | *An early decline in eGFR with finerenone is expected, but unlike placebo, it is not associated with worse outcomes and should not warrant discontinuation of therapy* |
|  | Shah et. Al. 2024 | Multicenter | 1145 | 575 | 570 | 72 | 12 | 298 | 18 | 282 | 12 | 57.5 | 2.5 | 57.5 | 2.5 | Semaglutide | 573 | Placebo | 573 | Moderate | In obesity-related HFpEF, semaglutide significantly improved symptoms, functional capacity, and reduced diuretic use compared with placebo over 52 weeks. |
|  | Peikert et. Al. 2024 | Multicenter | 6263 | 3516 | 2747 | 71.7 | 27.6 | N/A | N/A | N/A | N/A | 54.2 | 8.8 | 56 | 9.2 | Beta-blocker | 5177 | Placebo | 1086 | High | Dapagliflozin reduced the risk of cardiovascular death or worsening HF in patients with HFmrEF/HFpEF, irrespective of beta-blocker use, with consistent safety |
|  | Lewis et. Al. 2021 | Multicenter | 94 | 51 | 43 | 78 | 12 | 286 | 160 | 262 | 173 | 67 | 7 | 65 | 10 | Pirfenidone | 47 | Placebo | 47 | Moderate | *Pirfenidone reduced myocardial fibrosis (ECV) and NT-proBNP compared with placebo in HFpEF patients with myocardial fibrosis after 52 weeks* |
|  | Komajda et. al. 2017 | Multicenter | 179 | 63 | 116 | 72 | 18 | 323 | 244 | 321 | 257 | 60 | 6 | 61 | 6 | Ivabradine | 95 | Placebo | 84 | Low | Ivabradine reduced heart rate but did not improve symptoms, exercise capacity, or biomarkers in HFpEF |

Figure S1. Ranking of All Cause Mortality in the given study

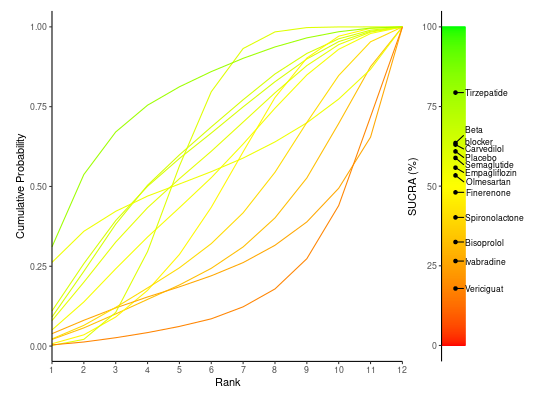

Table S2. Comparison Ranking of each treatment in the given study

|  | Beta_blocker | Bisoprolol | Carvedilol | Empagliflozin | Finerenone | Ivabradine | Olmesartan | Placebo | Semaglutide | Spironolactone | Tirzepatide | Vericiguat |
| --- | --- | --- | --- | --- | --- | --- | --- | --- | --- | --- | --- | --- |
| Beta_blocker | Beta_blocker | 2.56 (0.2, 33.38) | 1 (0.09, 11.12) | 1.13 (0.01, 75.17) | 1.46 (0.22, 9.87) | 4.17 (0.16, 222.73) | 1.31 (0.13, 13.73) | 1.13 (0.22, 5.93) | 1.13 (0.1, 12.04) | 1.89 (0.18, 19.91) | 0.57 (0.05, 6.51) | 4.47 (0.38, 53.43) |
| Bisoprolol | 0.39 (0.03, 5.01) | Bisoprolol | 0.39 (0.03, 5.2) | 0.43 (0.01, 33.25) | 0.57 (0.07, 4.94) | 1.63 (0.05, 99.39) | 0.51 (0.04, 6.71) | 0.44 (0.06, 3.12) | 0.44 (0.03, 5.83) | 0.73 (0.06, 9.53) | 0.22 (0.02, 3.18) | 1.75 (0.12, 25.56) |
| Carvedilol | 1 (0.09, 11.25) | 2.57 (0.19, 35.9) | Carvedilol | 1.13 (0.01, 79.85) | 1.47 (0.2, 11.02) | 4.18 (0.15, 237.46) | 1.32 (0.12, 14.95) | 1.14 (0.2, 6.59) | 1.14 (0.1, 13.11) | 1.89 (0.17, 21.43) | 0.58 (0.05, 6.93) | 4.5 (0.36, 56.76) |
| Empagliflozin | 0.88 (0.01, 68.16) | 2.31 (0.03, 195.85) | 0.89 (0.01, 70.69) | Empagliflozin | 1.3 (0.02, 80.27) | 3.91 (0.03, 824.96) | 1.17 (0.02, 90.28) | 1.01 (0.02, 56.57) | 0.99 (0.02, 80.8) | 1.68 (0.03, 129.15) | 0.52 (0.01, 40.89) | 4.02 (0.06, 330.11) |
| Finerenone | 0.68 (0.1, 4.54) | 1.76 (0.2, 15.18) | 0.68 (0.09, 5) | 0.77 (0.01, 41.18) | Finerenone | 2.79 (0.15, 127.14) | 0.9 (0.13, 6.01) | 0.77 (0.3, 2) | 0.77 (0.11, 5.41) | 1.29 (0.19, 8.71) | 0.39 (0.05, 2.93) | 3.07 (0.39, 24.22) |
| Ivabradine | 0.24 (0, 6.43) | 0.62 (0.01, 19.72) | 0.24 (0, 6.61) | 0.26 (0, 33.13) | 0.36 (0.01, 6.89) | Ivabradine | 0.31 (0.01, 8.51) | 0.28 (0.01, 4.61) | 0.27 (0, 7.4) | 0.45 (0.01, 12.18) | 0.14 (0, 3.87) | 1.08 (0.02, 31.13) |
| Olmesartan | 0.76 (0.07, 7.88) | 1.97 (0.15, 25.01) | 0.76 (0.07, 8.42) | 0.85 (0.01, 55.74) | 1.11 (0.17, 7.62) | 3.18 (0.12, 170.74) | Olmesartan | 0.86 (0.16, 4.51) | 0.86 (0.08, 9.29) | 1.44 (0.13, 15.05) | 0.44 (0.04, 4.97) | 3.42 (0.29, 40.6) |
| Placebo | 0.88 (0.17, 4.56) | 2.26 (0.32, 16.14) | 0.88 (0.15, 5.04) | 0.99 (0.02, 48.27) | 1.29 (0.5, 3.34) | 3.58 (0.22, 143.93) | 1.16 (0.22, 6.19) | Placebo | 0.99 (0.18, 5.54) | 1.67 (0.31, 8.87) | 0.51 (0.09, 2.98) | 3.95 (0.64, 24.82) |
| Semaglutide | 0.89 (0.08, 9.58) | 2.29 (0.17, 30.85) | 0.88 (0.08, 10.3) | 1.01 (0.01, 65.79) | 1.3 (0.18, 9.34) | 3.69 (0.14, 205.82) | 1.17 (0.11, 12.93) | 1.01 (0.18, 5.61) | Semaglutide | 1.68 (0.15, 18.51) | 0.51 (0.04, 5.99) | 3.99 (0.33, 48.82) |
| Spironolactone | 0.53 (0.05, 5.57) | 1.36 (0.1, 17.99) | 0.53 (0.05, 5.95) | 0.59 (0.01, 39.41) | 0.77 (0.11, 5.35) | 2.21 (0.08, 123.37) | 0.7 (0.07, 7.52) | 0.6 (0.11, 3.22) | 0.6 (0.05, 6.71) | Spironolactone | 0.3 (0.03, 3.58) | 2.37 (0.2, 28.76) |
| Tirzepatide | 1.75 (0.15, 19.79) | 4.46 (0.31, 62.91) | 1.73 (0.14, 20.91) | 1.94 (0.02, 139.36) | 2.54 (0.34, 18.94) | 7.27 (0.26, 416.43) | 2.29 (0.2, 25.58) | 1.97 (0.34, 11.6) | 1.96 (0.17, 23.04) | 3.28 (0.28, 38.06) | Tirzepatide | 7.8 (0.61, 100.23) |
| Vericiguat | 0.22 (0.02, 2.62) | 0.57 (0.04, 8.22) | 0.22 (0.02, 2.78) | 0.25 (0, 17.76) | 0.33 (0.04, 2.56) | 0.93 (0.03, 54.44) | 0.29 (0.02, 3.49) | 0.25 (0.04, 1.56) | 0.25 (0.02, 3.06) | 0.42 (0.03, 4.96) | 0.13 (0.01, 1.64) | Vericiguat |

Figure S2. Ranking of Cardiovascular Mortality

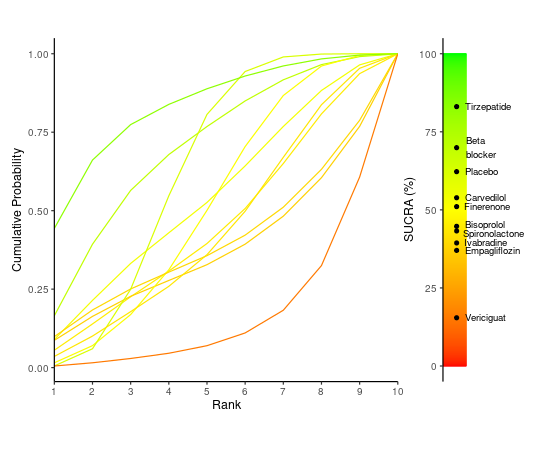

Table S3. Comparison of all treatments for Cardiovascular Mortality

|  | Beta_blocker | Bisoprolol | Carvedilol | Empagliflozin | Finerenone | Ivabradine | Placebo | Spironolactone | Tirzepatide | Vericiguat |
| --- | --- | --- | --- | --- | --- | --- | --- | --- | --- | --- |
| Beta_blocker | Beta_blocker | 2.24 (0.12, 39.12) | 1.64 (0.09, 29.43) | 3.47 (0.09, 249.86) | 1.73 (0.19, 16.1) | 3.1 (0.08, 231.33) | 1.36 (0.2, 9.32) | 2.25 (0.15, 34.55) | 0.57 (0.03, 9.43) | 7.83 (0.45, 145.16) |
| Bisoprolol | 0.45 (0.03, 8.12) | Bisoprolol | 0.73 (0.04, 15.7) | 1.56 (0.03, 125.02) | 0.78 (0.07, 8.67) | 1.39 (0.03, 113.8) | 0.61 (0.07, 5.26) | 1.02 (0.06, 18.21) | 0.26 (0.01, 5.03) | 3.54 (0.17, 77.95) |
| Carvedilol | 0.61 (0.03, 10.97) | 1.37 (0.06, 28.33) | Carvedilol | 2.1 (0.05, 170.13) | 1.05 (0.09, 11.82) | 1.9 (0.04, 146.71) | 0.83 (0.1, 7.21) | 1.38 (0.08, 23.93) | 0.35 (0.02, 6.74) | 4.82 (0.23, 103.68) |
| Empagliflozin | 0.29 (0, 11.56) | 0.64 (0.01, 29.5) | 0.48 (0.01, 21.8) | Empagliflozin | 0.5 (0.01, 14.45) | 0.91 (0.01, 129.29) | 0.4 (0.01, 9.63) | 0.65 (0.01, 27.07) | 0.16 (0, 7.23) | 2.27 (0.03, 110.28) |
| Finerenone | 0.58 (0.06, 5.15) | 1.29 (0.12, 14.39) | 0.95 (0.08, 10.88) | 1.99 (0.07, 104.93) | Finerenone | 1.78 (0.06, 99.04) | 0.79 (0.26, 2.32) | 1.3 (0.14, 11.98) | 0.33 (0.03, 3.37) | 4.54 (0.41, 53.78) |
| Ivabradine | 0.32 (0, 12.74) | 0.72 (0.01, 32.87) | 0.53 (0.01, 23.8) | 1.1 (0.01, 159.44) | 0.56 (0.01, 15.62) | Ivabradine | 0.45 (0.01, 10.48) | 0.73 (0.01, 30.39) | 0.18 (0, 8.01) | 2.53 (0.03, 124.27) |
| Placebo | 0.74 (0.11, 4.94) | 1.63 (0.19, 14.12) | 1.2 (0.14, 10.48) | 2.51 (0.1, 116.27) | 1.27 (0.43, 3.79) | 2.24 (0.1, 109.04) | Placebo | 1.66 (0.24, 11.44) | 0.42 (0.05, 3.24) | 5.74 (0.67, 53.78) |
| Spironolactone | 0.44 (0.03, 6.86) | 0.98 (0.05, 17.97) | 0.72 (0.04, 12.8) | 1.54 (0.04, 105.54) | 0.77 (0.08, 7.08) | 1.37 (0.03, 99.85) | 0.6 (0.09, 4.17) | Spironolactone | 0.25 (0.01, 4.11) | 3.5 (0.19, 66.58) |
| Tirzepatide | 1.76 (0.11, 29.91) | 3.91 (0.2, 76.77) | 2.87 (0.15, 57.88) | 6.12 (0.14, 455.26) | 3.03 (0.3, 31.97) | 5.51 (0.12, 427.88) | 2.39 (0.31, 19.16) | 3.97 (0.24, 67.66) | Tirzepatide | 13.96 (0.72, 287.52) |
| Vericiguat | 0.13 (0.01, 2.24) | 0.28 (0.01, 5.92) | 0.21 (0.01, 4.41) | 0.44 (0.01, 35.11) | 0.22 (0.02, 2.44) | 0.4 (0.01, 31.96) | 0.17 (0.02, 1.48) | 0.29 (0.02, 5.15) | 0.07 (0, 1.4) | Vericiguat |

Figure S4. Rehospitalization SUCRA Ranking

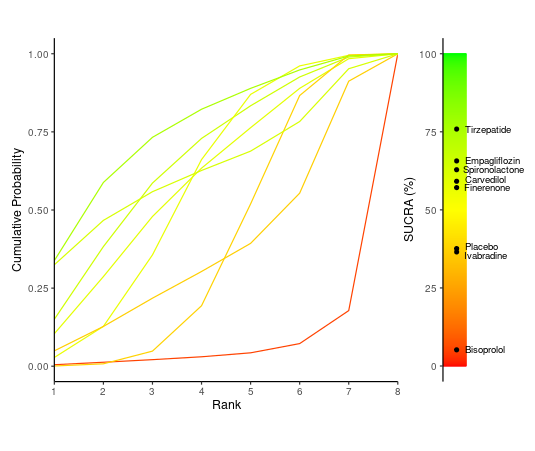

Table S4. Comparison of all Treatments for Rehospitalization

|  | Bisoprolol | Carvedilol | Empagliflozin | Finerenone | Ivabradine | Placebo | Spironolactone | Tirzepatide |
| --- | --- | --- | --- | --- | --- | --- | --- | --- |
| Bisoprolol | Bisoprolol | 0.16 (0.01, 1.36) | 0.14 (0.01, 1.23) | 0.17 (0.02, 1.2) | 0.24 (0.02, 2.89) | 0.21 (0.02, 1.3) | 0.13 (0.01, 1.92) | 0.11 (0.01, 1.04) |
| Carvedilol | 6.41 (0.73, 83.56) | Carvedilol | 0.89 (0.16, 4.64) | 1.07 (0.26, 4.2) | 1.59 (0.24, 9.66) | 1.32 (0.4, 4.22) | 0.83 (0.06, 8.48) | 0.69 (0.11, 3.98) |
| Empagliflozin | 7.3 (0.81, 93.24) | 1.13 (0.22, 6.08) | Empagliflozin | 1.2 (0.3, 4.9) | 1.8 (0.27, 11.05) | 1.48 (0.46, 4.87) | 0.93 (0.07, 9.85) | 0.78 (0.13, 4.64) |
| Finerenone | 5.95 (0.84, 64) | 0.94 (0.24, 3.83) | 0.83 (0.2, 3.36) | Finerenone | 1.49 (0.29, 7.08) | 1.23 (0.58, 2.61) | 0.79 (0.07, 6.59) | 0.65 (0.14, 2.86) |
| Ivabradine | 4.2 (0.35, 60.24) | 0.63 (0.1, 4.15) | 0.56 (0.09, 3.69) | 0.67 (0.14, 3.46) | Ivabradine | 0.83 (0.2, 3.57) | 0.52 (0.03, 6.42) | 0.43 (0.06, 3.08) |
| Placebo | 4.83 (0.77, 46.27) | 0.76 (0.24, 2.47) | 0.67 (0.21, 2.19) | 0.81 (0.38, 1.73) | 1.21 (0.28, 4.94) | Placebo | 0.64 (0.06, 4.76) | 0.53 (0.13, 1.95) |
| Spironolactone | 7.96 (0.52, 183.94) | 1.2 (0.12, 16.49) | 1.07 (0.1, 13.98) | 1.27 (0.15, 15.34) | 1.91 (0.16, 29.86) | 1.57 (0.21, 17.19) | Spironolactone | 0.8 (0.07, 12.43) |
| Tirzepatide | 9.47 (0.96, 141.59) | 1.46 (0.25, 8.99) | 1.28 (0.22, 7.78) | 1.54 (0.35, 7.4) | 2.3 (0.32, 16.57) | 1.9 (0.51, 7.68) | 1.25 (0.08, 13.38) | Tirzepatide |

Figure S5. Ranking of KCC Quality of Life

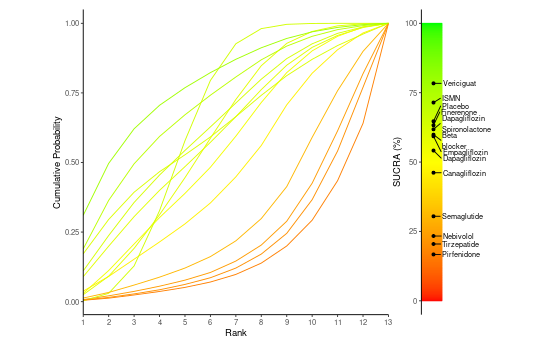

Table S5. Comparison of all Treatments for KCC Quality of Life.

|  | Beta_blocker | Canagliflozin | Dapagliflozin | Dapagliflozin_Spironolactone | Empagliflozin | Finerenone | ISMN | Nebivolol | Pirfenidone | Placebo | Semaglutide | Tirzepatide | Vericiguat |
| --- | --- | --- | --- | --- | --- | --- | --- | --- | --- | --- | --- | --- | --- |
| Beta_blocker | Beta_blocker | 2.84 (-16.59, 22.03) | 1.07 (-18.15, 20.22) | -0.61 (-24.06, 23.05) | 0.21 (-15.54, 16.03) | -0.7 (-19.92, 18.62) | -2.47 (-21.78, 16.79) | 8.49 (-10.89, 27.83) | 10.91 (-9.02, 30.68) | -0.49 (-14.05, 13.1) | 6.5 (-12.9, 25.86) | 9.36 (-9.9, 28.82) | -4.17 (-23.61, 15.26) |
| Canagliflozin | -2.84 (-22.03, 16.59) | Canagliflozin | -1.76 (-20.95, 17.63) | -3.43 (-27.01, 20.21) | -2.59 (-18.45, 13.38) | -3.5 (-22.74, 15.9) | -5.28 (-24.64, 13.83) | 5.65 (-13.77, 25.2) | 8.12 (-11.72, 27.8) | -3.34 (-16.96, 10.39) | 3.68 (-15.69, 23.02) | 6.56 (-12.77, 25.76) | -6.96 (-26.28, 12.33) |
| Dapagliflozin | -1.07 (-20.22, 18.15) | 1.76 (-17.63, 20.95) | Dapagliflozin | -1.7 (-15.36, 12.06) | -0.85 (-16.49, 14.79) | -1.8 (-20.9, 17.76) | -3.57 (-22.84, 15.85) | 7.42 (-11.97, 26.83) | 9.82 (-9.95, 29.6) | -1.58 (-15.25, 11.84) | 5.42 (-14.03, 24.7) | 8.23 (-10.88, 27.56) | -5.2 (-24.53, 14.03) |
| Dapagliflozin_Spironolactone | 0.61 (-23.05, 24.06) | 3.43 (-20.21, 27.01) | 1.7 (-12.06, 15.36) | Dapagliflozin_Spironolactone | 0.85 (-20.1, 21.46) | -0.09 (-23.65, 23.53) | -1.82 (-25.51, 21.72) | 9.03 (-14.64, 32.62) | 11.53 (-12.46, 35.45) | 0.12 (-19.24, 19.23) | 7.09 (-16.55, 30.63) | 9.94 (-13.77, 33.77) | -3.54 (-27.13, 19.95) |
| Empagliflozin | -0.21 (-16.03, 15.54) | 2.59 (-13.38, 18.45) | 0.85 (-14.79, 16.49) | -0.85 (-21.46, 20.1) | Empagliflozin | -0.94 (-16.7, 14.93) | -2.68 (-18.54, 13.07) | 8.24 (-7.75, 24.2) | 10.7 (-5.87, 27.06) | -0.74 (-8.59, 7.14) | 6.28 (-9.49, 22) | 9.14 (-6.8, 24.78) | -4.39 (-20.18, 11.4) |
| Finerenone | 0.7 (-18.62, 19.92) | 3.5 (-15.9, 22.74) | 1.8 (-17.76, 20.9) | 0.09 (-23.53, 23.65) | 0.94 (-14.93, 16.7) | Finerenone | -1.76 (-21.19, 17.38) | 9.17 (-10.5, 28.61) | 11.64 (-8.39, 31.34) | 0.19 (-13.6, 13.84) | 7.19 (-12.16, 26.3) | 10.04 (-9.39, 29.2) | -3.47 (-23.04, 16.01) |
| ISMN | 2.47 (-16.79, 21.78) | 5.28 (-13.83, 24.64) | 3.57 (-15.85, 22.84) | 1.82 (-21.72, 25.51) | 2.68 (-13.07, 18.54) | 1.76 (-17.38, 21.19) | ISMN | 10.96 (-8.36, 30.46) | 13.4 (-6.62, 33.1) | 1.98 (-11.66, 15.7) | 8.97 (-10.41, 28.26) | 11.81 (-7.61, 31.3) | -1.72 (-21.06, 17.84) |
| Nebivolol | -8.49 (-27.83, 10.89) | -5.65 (-25.2, 13.77) | -7.42 (-26.83, 11.97) | -9.03 (-32.62, 14.64) | -8.24 (-24.2, 7.75) | -9.17 (-28.61, 10.5) | -10.96 (-30.46, 8.36) | Nebivolol | 2.43 (-17.54, 22.37) | -8.98 (-22.65, 4.82) | -1.99 (-21.42, 17.53) | 0.86 (-18.75, 20.45) | -12.64 (-31.92, 6.8) |
| Pirfenidone | -10.91 (-30.68, 9.02) | -8.12 (-27.8, 11.72) | -9.82 (-29.6, 9.95) | -11.53 (-35.45, 12.46) | -10.7 (-27.06, 5.87) | -11.64 (-31.34, 8.39) | -13.4 (-33.1, 6.62) | -2.43 (-22.37, 17.54) | Pirfenidone | -11.39 (-25.84, 3.09) | -4.45 (-24.41, 15.65) | -1.53 (-21.45, 18.41) | -15.05 (-34.82, 4.81) |
| Placebo | 0.49 (-13.1, 14.05) | 3.34 (-10.39, 16.96) | 1.58 (-11.84, 15.25) | -0.12 (-19.23, 19.24) | 0.74 (-7.14, 8.59) | -0.19 (-13.84, 13.6) | -1.98 (-15.7, 11.66) | 8.98 (-4.82, 22.65) | 11.39 (-3.09, 25.84) | Placebo | 7.02 (-6.66, 20.65) | 9.88 (-3.85, 23.57) | -3.65 (-17.37, 10.04) |
| Semaglutide | -6.5 (-25.86, 12.9) | -3.68 (-23.02, 15.69) | -5.42 (-24.7, 14.03) | -7.09 (-30.63, 16.55) | -6.28 (-22, 9.49) | -7.19 (-26.3, 12.16) | -8.97 (-28.26, 10.41) | 1.99 (-17.53, 21.42) | 4.45 (-15.65, 24.41) | -7.02 (-20.65, 6.66) | Semaglutide | 2.89 (-16.48, 22.49) | -10.65 (-29.99, 8.81) |
| Tirzepatide | -9.36 (-28.82, 9.9) | -6.56 (-25.76, 12.77) | -8.23 (-27.56, 10.88) | -9.94 (-33.77, 13.77) | -9.14 (-24.78, 6.8) | -10.04 (-29.2, 9.39) | -11.81 (-31.3, 7.61) | -0.86 (-20.45, 18.75) | 1.53 (-18.41, 21.45) | -9.88 (-23.57, 3.85) | -2.89 (-22.49, 16.48) | Tirzepatide | -13.5 (-32.87, 6.02) |
| Vericiguat | 4.17 (-15.26, 23.61) | 6.96 (-12.33, 26.28) | 5.2 (-14.03, 24.53) | 3.54 (-19.95, 27.13) | 4.39 (-11.4, 20.18) | 3.47 (-16.01, 23.04) | 1.72 (-17.84, 21.06) | 12.64 (-6.8, 31.92) | 15.05 (-4.81, 34.82) | 3.65 (-10.04, 17.37) | 10.65 (-8.81, 29.99) | 13.5 (-6.02, 32.87) | Vericiguat |

Figure S6. SUCRA Ranking for 6 Minute Walking Test.

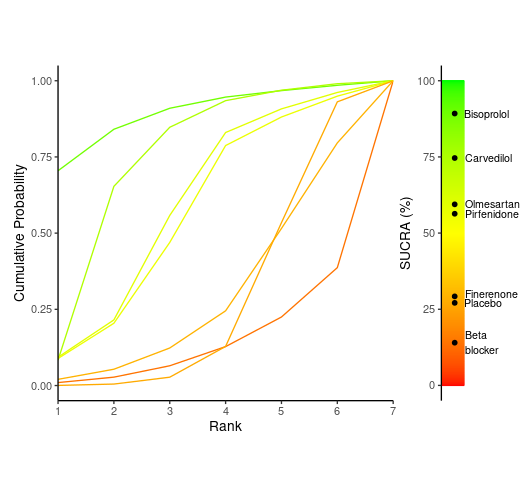

Table S6. Comparison for all treatments for 6 Minute Walking Test

|  | Beta_blocker | Bisoprolol | Carvedilol | Finerenone | Olmesartan | Pirfenidone | Placebo |
| --- | --- | --- | --- | --- | --- | --- | --- |
| Beta_blocker | Beta_blocker | -27.88 (-62.84, 7.16) | -20.91 (-49.63, 7.6) | -4.99 (-33.65, 23.83) | -14.96 (-43.45, 13.93) | -13.95 (-43.12, 15.37) | -5.01 (-25.2, 15.66) |
| Bisoprolol | 27.88 (-7.16, 62.84) | Bisoprolol | 7 (-13.11, 27.16) | 22.93 (-12.02, 58.1) | 12.96 (-21.96, 47.91) | 14.01 (-21.36, 49.16) | 22.95 (-5.68, 51.44) |
| Carvedilol | 20.91 (-7.6, 49.63) | -7 (-27.16, 13.11) | Carvedilol | 15.95 (-12.5, 44.84) | 5.95 (-22.46, 34.5) | 7.04 (-21.91, 36.33) | 15.96 (-4.22, 36.18) |
| Finerenone | 4.99 (-23.83, 33.65) | -22.93 (-58.1, 12.02) | -15.95 (-44.84, 12.5) | Finerenone | -9.98 (-38.47, 18.82) | -8.96 (-38.05, 20.4) | 0.01 (-20.38, 20.13) |
| Olmesartan | 14.96 (-13.93, 43.45) | -12.96 (-47.91, 21.96) | -5.95 (-34.5, 22.46) | 9.98 (-18.82, 38.47) | Olmesartan | 1.05 (-28.36, 30.1) | 10 (-10.45, 30.21) |
| Pirfenidone | 13.95 (-15.37, 43.12) | -14.01 (-49.16, 21.36) | -7.04 (-36.33, 21.91) | 8.96 (-20.4, 38.05) | -1.05 (-30.1, 28.36) | Pirfenidone | 8.99 (-12.12, 30.05) |
| Placebo | 5.01 (-15.66, 25.2) | -22.95 (-51.44, 5.68) | -15.96 (-36.18, 4.22) | -0.01 (-20.13, 20.38) | -10 (-30.21, 10.45) | -8.99 (-30.05, 12.12) | Placebo |

Figure S7. SUCRA for NT-proBNP

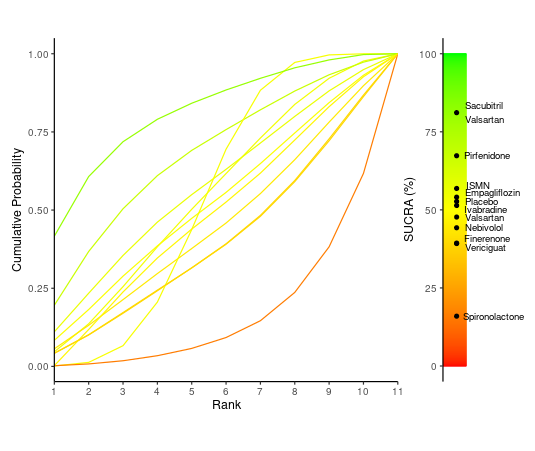

Table S7. Comparison for all treatments for NT-proBNP

|  | Empagliflozin | Finerenone | ISMN | Ivabradine | Nebivolol | Pirfenidone | Placebo | Sacubitril_Valsartan | Spironolactone | Valsartan | Vericiguat |
| --- | --- | --- | --- | --- | --- | --- | --- | --- | --- | --- | --- |
| Empagliflozin | Empagliflozin | 78.03 (-406.66, 557.71) | -18.36 (-505.39, 461.9) | 10.82 (-476.66, 497.03) | 50.57 (-434.45, 534.82) | -80.11 (-561.65, 404.22) | 8.88 (-269.02, 288.94) | -168.88 (-685.22, 346.96) | 208.93 (-181.16, 602.77) | 19.86 (-464.08, 505.29) | 80.49 (-407.98, 566.3) |
| Finerenone | -78.03 (-557.71, 406.66) | Finerenone | -96.63 (-656.25, 460.13) | -67.77 (-620.83, 487.77) | -27.88 (-583.32, 528.58) | -157.57 (-712.45, 400.4) | -69.54 (-460.86, 323.61) | -245.7 (-829.71, 333.4) | 131.7 (-347.87, 613.94) | -58.4 (-617.45, 495.99) | 0.98 (-554.19, 559.37) |
| ISMN | 18.36 (-461.9, 505.39) | 96.63 (-460.13, 656.25) | ISMN | 30.15 (-523.76, 586.95) | 69.86 (-493.29, 626.87) | -60.31 (-614.09, 498.42) | 27.34 (-364.33, 424.54) | -149.87 (-735.06, 432.41) | 229.18 (-252.97, 712.87) | 38.65 (-518.12, 593.29) | 99.63 (-456.8, 658.35) |
| Ivabradine | -10.82 (-497.03, 476.66) | 67.77 (-487.77, 620.83) | -30.15 (-586.95, 523.76) | Ivabradine | 38.64 (-515.52, 591.55) | -91.57 (-648.59, 472.13) | -2.59 (-395.62, 389.81) | -180.56 (-763.26, 403.81) | 198.39 (-281.74, 685.14) | 9.04 (-549.22, 571.04) | 68.34 (-489.06, 629.86) |
| Nebivolol | -50.57 (-534.82, 434.45) | 27.88 (-528.58, 583.32) | -69.86 (-626.87, 493.29) | -38.64 (-591.55, 515.52) | Nebivolol | -131.45 (-689.09, 432.26) | -42.04 (-437.35, 354.6) | -219.63 (-805.71, 367.94) | 158.61 (-323.74, 646.95) | -30.35 (-584.93, 527.4) | 29.22 (-531.92, 585.99) |
| Pirfenidone | 80.11 (-404.22, 561.65) | 157.57 (-400.4, 712.45) | 60.31 (-498.42, 614.09) | 91.57 (-472.13, 648.59) | 131.45 (-432.26, 689.09) | Pirfenidone | 89.02 (-305.61, 482.65) | -89.04 (-678.11, 498.52) | 288.94 (-194.33, 772.22) | 100 (-460.72, 657.7) | 159.45 (-402.27, 721.14) |
| Placebo | -8.88 (-288.94, 269.02) | 69.54 (-323.61, 460.86) | -27.34 (-424.54, 364.33) | 2.59 (-389.81, 395.62) | 42.04 (-354.6, 437.35) | -89.02 (-482.65, 305.61) | Placebo | -177.05 (-608.95, 255.76) | 201.5 (-74.61, 480.72) | 11.26 (-385.41, 407.34) | 70.7 (-324.82, 468.02) |
| Sacubitril_Valsartan | 168.88 (-346.96, 685.22) | 245.7 (-333.4, 829.71) | 149.87 (-432.41, 735.06) | 180.56 (-403.81, 763.26) | 219.63 (-367.94, 805.71) | 89.04 (-498.52, 678.11) | 177.05 (-255.76, 608.95) | Sacubitril_Valsartan | 377.44 (-136.53, 894.43) | 188.66 (12.24, 367.94) | 247.48 (-337.12, 837.07) |
| Spironolactone | -208.93 (-602.77, 181.16) | -131.7 (-613.94, 347.87) | -229.18 (-712.87, 252.97) | -198.39 (-685.14, 281.74) | -158.61 (-646.95, 323.74) | -288.94 (-772.22, 194.33) | -201.5 (-480.72, 74.61) | -377.44 (-894.43, 136.53) | Spironolactone | -189.46 (-675.75, 294.8) | -129.78 (-613.37, 357.14) |
| Valsartan | -19.86 (-505.29, 464.08) | 58.4 (-495.99, 617.45) | -38.65 (-593.29, 518.12) | -9.04 (-571.04, 549.22) | 30.35 (-527.4, 584.93) | -100 (-657.7, 460.72) | -11.26 (-407.34, 385.41) | -188.66 (-367.94, -12.24) | 189.46 (-294.8, 675.75) | Valsartan | 59.44 (-496.15, 616.55) |
| Vericiguat | -80.49 (-566.3, 407.98) | -0.98 (-559.37, 554.19) | -99.63 (-658.35, 456.8) | -68.34 (-629.86, 489.06) | -29.22 (-585.99, 531.92) | -159.45 (-721.14, 402.27) | -70.7 (-468.02, 324.82) | -247.48 (-837.07, 337.12) | 129.78 (-357.14, 613.37) | -59.44 (-616.55, 496.15) | Vericiguat |

Figure S8. SUCRA for EGFR

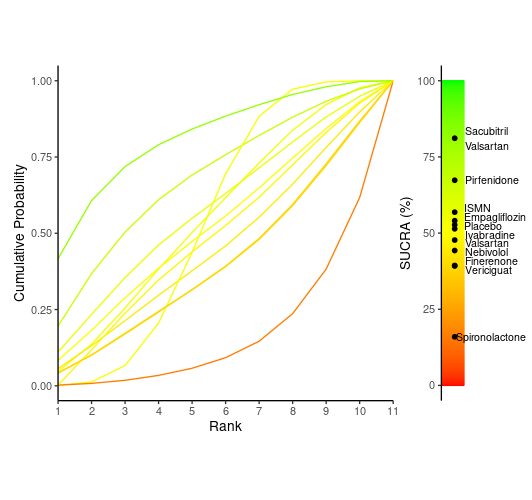

Table S8. Comparison for all treatments for EGFR

|  | Empagliflozin | Finerenone | ISMN | Ivabradine | Nebivolol | Pirfenidone | Placebo | Sacubitril_Valsartan | Spironolactone | Valsartan | Vericiguat |
| --- | --- | --- | --- | --- | --- | --- | --- | --- | --- | --- | --- |
| Empagliflozin | Empagliflozin | 78.03 (-406.66, 557.71) | -18.36 (-505.39, 461.9) | 10.82 (-476.66, 497.03) | 50.57 (-434.45, 534.82) | -80.11 (-561.65, 404.22) | 8.88 (-269.02, 288.94) | -168.88 (-685.22, 346.96) | 208.93 (-181.16, 602.77) | 19.86 (-464.08, 505.29) | 80.49 (-407.98, 566.3) |
| Finerenone | -78.03 (-557.71, 406.66) | Finerenone | -96.63 (-656.25, 460.13) | -67.77 (-620.83, 487.77) | -27.88 (-583.32, 528.58) | -157.57 (-712.45, 400.4) | -69.54 (-460.86, 323.61) | -245.7 (-829.71, 333.4) | 131.7 (-347.87, 613.94) | -58.4 (-617.45, 495.99) | 0.98 (-554.19, 559.37) |
| ISMN | 18.36 (-461.9, 505.39) | 96.63 (-460.13, 656.25) | ISMN | 30.15 (-523.76, 586.95) | 69.86 (-493.29, 626.87) | -60.31 (-614.09, 498.42) | 27.34 (-364.33, 424.54) | -149.87 (-735.06, 432.41) | 229.18 (-252.97, 712.87) | 38.65 (-518.12, 593.29) | 99.63 (-456.8, 658.35) |
| Ivabradine | -10.82 (-497.03, 476.66) | 67.77 (-487.77, 620.83) | -30.15 (-586.95, 523.76) | Ivabradine | 38.64 (-515.52, 591.55) | -91.57 (-648.59, 472.13) | -2.59 (-395.62, 389.81) | -180.56 (-763.26, 403.81) | 198.39 (-281.74, 685.14) | 9.04 (-549.22, 571.04) | 68.34 (-489.06, 629.86) |
| Nebivolol | -50.57 (-534.82, 434.45) | 27.88 (-528.58, 583.32) | -69.86 (-626.87, 493.29) | -38.64 (-591.55, 515.52) | Nebivolol | -131.45 (-689.09, 432.26) | -42.04 (-437.35, 354.6) | -219.63 (-805.71, 367.94) | 158.61 (-323.74, 646.95) | -30.35 (-584.93, 527.4) | 29.22 (-531.92, 585.99) |
| Pirfenidone | 80.11 (-404.22, 561.65) | 157.57 (-400.4, 712.45) | 60.31 (-498.42, 614.09) | 91.57 (-472.13, 648.59) | 131.45 (-432.26, 689.09) | Pirfenidone | 89.02 (-305.61, 482.65) | -89.04 (-678.11, 498.52) | 288.94 (-194.33, 772.22) | 100 (-460.72, 657.7) | 159.45 (-402.27, 721.14) |
| Placebo | -8.88 (-288.94, 269.02) | 69.54 (-323.61, 460.86) | -27.34 (-424.54, 364.33) | 2.59 (-389.81, 395.62) | 42.04 (-354.6, 437.35) | -89.02 (-482.65, 305.61) | Placebo | -177.05 (-608.95, 255.76) | 201.5 (-74.61, 480.72) | 11.26 (-385.41, 407.34) | 70.7 (-324.82, 468.02) |
| Sacubitril_Valsartan | 168.88 (-346.96, 685.22) | 245.7 (-333.4, 829.71) | 149.87 (-432.41, 735.06) | 180.56 (-403.81, 763.26) | 219.63 (-367.94, 805.71) | 89.04 (-498.52, 678.11) | 177.05 (-255.76, 608.95) | Sacubitril_Valsartan | 377.44 (-136.53, 894.43) | 188.66 (12.24, 367.94) | 247.48 (-337.12, 837.07) |
| Spironolactone | -208.93 (-602.77, 181.16) | -131.7 (-613.94, 347.87) | -229.18 (-712.87, 252.97) | -198.39 (-685.14, 281.74) | -158.61 (-646.95, 323.74) | -288.94 (-772.22, 194.33) | -201.5 (-480.72, 74.61) | -377.44 (-894.43, 136.53) | Spironolactone | -189.46 (-675.75, 294.8) | -129.78 (-613.37, 357.14) |
| Valsartan | -19.86 (-505.29, 464.08) | 58.4 (-495.99, 617.45) | -38.65 (-593.29, 518.12) | -9.04 (-571.04, 549.22) | 30.35 (-527.4, 584.93) | -100 (-657.7, 460.72) | -11.26 (-407.34, 385.41) | -188.66 (-367.94, -12.24) | 189.46 (-294.8, 675.75) | Valsartan | 59.44 (-496.15, 616.55) |
| Vericiguat | -80.49 (-566.3, 407.98) | -0.98 (-559.37, 554.19) | -99.63 (-658.35, 456.8) | -68.34 (-629.86, 489.06) | -29.22 (-585.99, 531.92) | -159.45 (-721.14, 402.27) | -70.7 (-468.02, 324.82) | -247.48 (-837.07, 337.12) | 129.78 (-357.14, 613.37) | -59.44 (-616.55, 496.15) | Vericiguat |

Risk of Bias

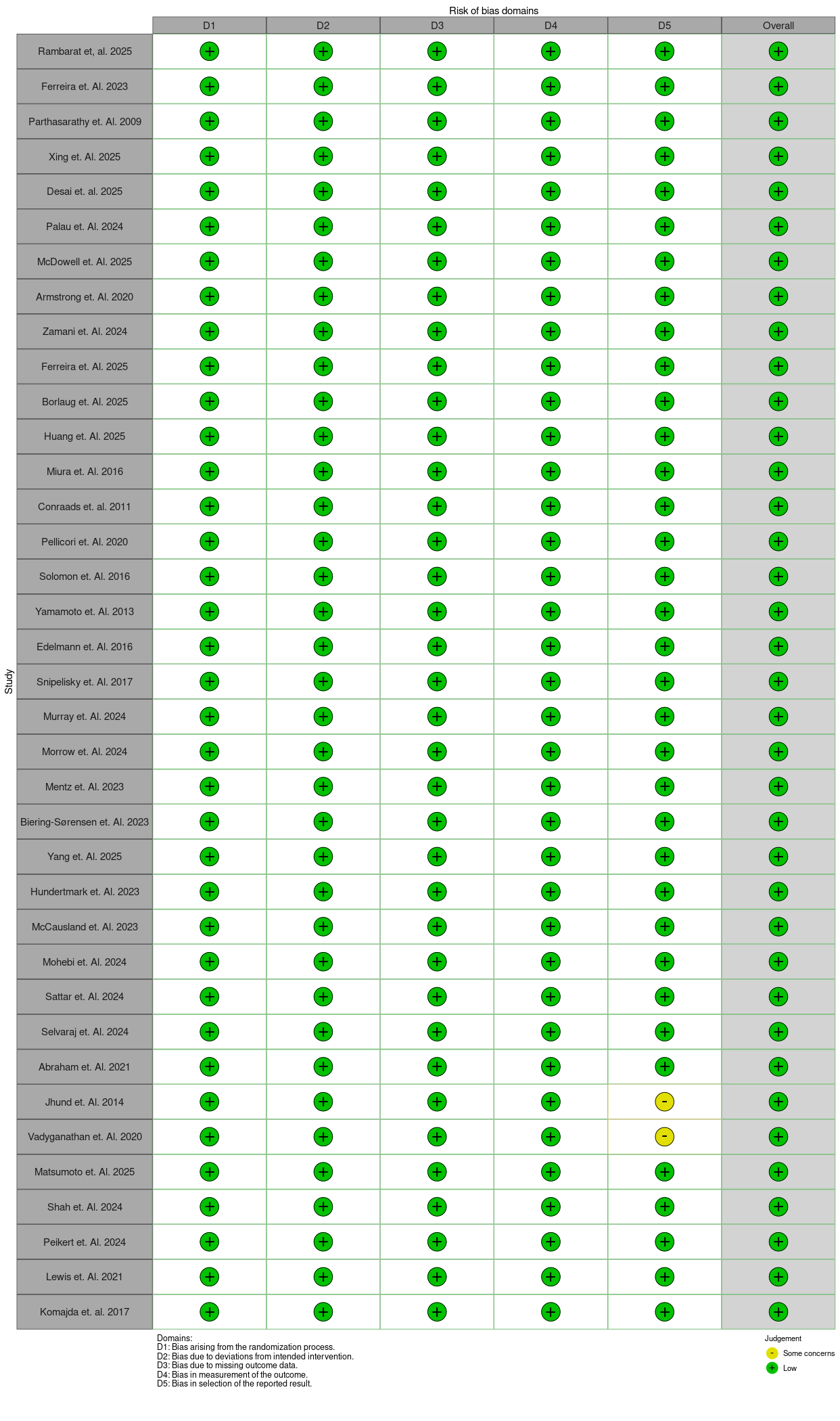
